# First-episode mild depression in young adults is a pre-proatherogenic condition even in the absence of subclinical metabolic syndrome: lowered lecithin-cholesterol acyltransferase as a key factor

**DOI:** 10.1101/2024.01.01.24300688

**Authors:** Michael Maes, Asara Vasupanrajit, Ketsupar Jirakran, Bo Zhou, Chavit Tunvirachaisakul, Abbas F. Almulla

**Author notes:** Joined first authorships (Michael Maes, Asara Vasupanrajit). joined corresponding authors. **Corresponding author:** Prof. Dr. Bo Zhou, M.D., Ph.D., Sichuan Provincial Center for Mental Health, Sichuan Provincial People’s Hospital, School of Medicine, University of Electronic Science and Technology of China, Chengdu 610072, China,; And; Dr. Abbas F. Almulla, Ph.D., Department of Psychiatry, Faculty of Medicine, Chulalongkorn University and King Chulalongkorn Memorial Hospital, the Thai Red Cross Society, Bangkok, Thailand.

## Abstract

**Background:** Major depression is classified into distinct subtypes: simple (SDMD) and major dysmood disorder (MDMD). MDMD patients exhibit elevated atherogenicity and decreased reverse cholesterol transport (RCT). However, comprehensive data regarding lipid metabolism is absent in first episode (FE)-SDMD.

**Objectives:** In this case-control study, plasma lipid levels, lecithin-cholesterol acyltransferase (LCAT), free cholesterol, ApoA1, ApoB, and ApoE are compared between academic students with first episode SDMD (FE-SDMD) (n=44) or SDMD (n=64) and control students (n=44), after excluding those with metabolic syndrome (MetS).

**Results:** LCAT is decreased, and free cholesterol and ApoE increased in subjects with SDMD and FE-SDMD as compared with controls. There were no significant alterations in high-density lipoprotein cholesterol (HDLc), apolipoprotein (Apo)A1, RCT, ApoB and triglycerides in SDMD. LCAT, free cholesterol and atherogenicity indices are significantly associated with suicidal behaviors and the SDMD phenome. The effects of LCAT on those phenome features is completely mediated by free cholesterol and brooding. SDMD and FE-SDMD patients without signs of subclinical MetS show lowered LCAT and increased free cholesterol as compared with normal controls. There are significant interactions between the SDMD and FE-SDMD diagnosis and subclinical MetS, which result in decreased HDLc and RCT, and an increased ApoB/ApoA ratio.

**Discussion:** FE-SDMD and SDMD are pre-proatherogenic states, because of decreased LCAT, and increased free cholesterol and ApoE, and their intersections with subclinical MetS. These aberrations may drive atherogenicity, and activation of peripheral and central oxidative, neuro-immune, and degenerative pathways. Individuals with FE-SDMD should be screened for increased atherogenicity risk by measuring LCAT, free cholesterol and ApoE.

## Introduction

1994 marked the first publication of the finding that major depressive disorder (MDD) is associated with decreased lecithin-cholesterol acyltransferase (LCAT) activity [27]. LCAT is a liver-secreted enzyme that exhibits a preferential affinity for free cholesterol contained within high-density lipoprotein (HDL) particles [1, 22, 24, 33, 39, 43, 52, 55, 66]. By facilitating the esterification of cholesterol on HDL particles, LCAT enhances the cholesterol transport pathway from cell membranes and peripheral tissues to HDL particles. This HDL core facilitates the transportation of cholesteryl esters to the liver, where they are catabolized and eliminated along with cholesterol. Because free cholesterol can diffuse directly into the arterial wall, induce cytotoxicity, inhibit the formation of membrane domains, accumulate, and crystallize in cells and macrophages, and trigger apoptotic pathways and cell death, free cholesterol contributes to atherosclerosis and the development of atheroma [58, 59]. Therefore, the process by which LCAT converts free cholesterol into cholesteryl esters is a critical element in the body’s elimination of the deleterious effects of free cholesterol [1]. In addition to HDL-cholesterol, apolipoprotein A1 (ApoA1) and paraoxonase 1 (PON1) are significant actors in this pathway referred to as “reverse cholesterol transport” (RCT) [1].

Both LCAT, ApoA1 and PON1 are components of HDL particles, and their anti-atherogenic, antioxidant, and anti-inflammatory properties are regulated by the HDL-ApoA-LCAT-PON1 complex [1, 8]. Furthermore, the cholesterol-accumulating ApoA1 is responsible for stimulating cholesterol esterification, activating LCAT, and incorporating cholesteryl esters into HDL; thus, it contributes to the maturation and formation of HDL [10, 20, 22, 68].

AT, and ApoA1 in comparison to healthy controls, according to a recent meta-analysis [1]. The results of the latter meta-analysis suggest that severe MDD is distinguished by a diminished RCT cycle activity and a reduced protective activity of the HDL-ApoA-LCAT-PON1 complex [1]. Significantly, a reduced LCAT index was also observed among first-degree relatives of MDD patients, providing evidence for the existence of a genetic element [27].

ApoE is an additional HDL-binding protein that, according to some sources [4, 8], may prevent lipid peroxidation and inflammation. However, there exists a significant correlation between plasma levels of ApoE and cholesterol, ApoB, triglycerides and low-density lipoprotein (LDL)-cholesterol [23, 57]. Furthermore, elevated levels of ApoE have the potential to serve as a prognostic indicator for cardiovascular disease and mortality, and Alzheimer’s disease [23, 60].

Moreover, MDD has been linked to elevated levels of proatherogenic lipids and lipoproteins, including ApoB, triglycerides, and LDL cholesterol [21, 27, 30, 37, 41, 53]. Probably the most reliable indicator of heightened atherogenicity in MDD is an elevated composite score based on plasma triglyceride, ApoB, LDLc, and free cholesterol levels [21]. Because each atherogenic lipoprotein, including LDL, very low-density lipoprotein, and intermediate-density lipoprotein, contains a single ApoB per particle, ApoB is an inclusive indicator of these lipoproteins [3, 56]. Therefore, a more accurate biomarker for MDD could be the ApoB/ApoA ratio [21, 35] or another ratio computed as atherogenicity / RCT; the latter being calculated as a composite using LCAT, HDL-cholesterol, and ApoA1 values [35].

However, three factors impede the interpretation atherogenicity and RCT data in samples of depression. First, Maes et al. [35] report that elevated atherogenicity indices and diminished RCT scores are exclusively observed in a subset of MDD patients. By utilizing machine learning methods, we identified two distinct populations of MDD patients—the first known as major dysmood disorder (MDMD) and the second as simple dysmood disorder (SDMD). In contrast to the latter subgroup, the former is distinguished by lowered levels of protective biomarkers (e.g., RCT, T regulatory cells, antioxidants, nerve growth factor), and increased levels of atherogenicity, immune activation, T cell activation, gut dysbiosis, and oxidative stress [34]. Patients with SDMD, in contrast, do not exhibit these disorders.

Second, MDMD patients have a greater recurrence of illness (ROI) index, which is inversely correlated with the RCT index and substantially and positively correlated with atherogenicity indices such as ApoB/ApoA [35]. The ROI index is calculated by adding the number of lifetime suicidal thoughts and attempts, and depression episodes together. Rephrased differently, ROI has a substantial impact on the RCT and atherogenicity outcomes; thus, failing to account for ROI may result in inaccurate outcomes [35].

Third, heightened atherogenicity and lowered RCT levels are potential indicators of metabolic syndrome (MetS); therefore, the existence of MetS in certain participants may serve as a robust predictor of the progression of atherogenicity and RCT results, rather than of MDD per se. MDD, MetS, and cardiovascular disorders (CVD) are strongly associated, which may be partially explained by MDD’s enhanced atherogenicity and decreased RCT [13, 27, 30, 37, 41]. Comorbidity with MetS in mood disorders has been linked to an increased frequency of episodes and suicidal ideation, a more complex affective presentation, and a decreased likelihood of recovery [13, 35, 36, 41, 65]. Additionally, it has been observed that individuals who have both MDD and MetS have significantly elevated atherogenicity indices compared to those with either disorder alone [13, 27, 30, 37, 41]. Conversely, within the combined MDD + MetS study group, no discernible correlations were found between MDD and lipid concentrations [21]. However, upon excluding MetS participants, it became evident that MDD is significantly correlated with (a) elevated levels of free cholesterol, triglycerides, ApoB, the Castelli risk index 1, and the ApoB/ApoA ratio; and (b) reduced HDL-cholesterol, ApoA1, and the RCT index.

However, no data is available regarding the presence of lipids in the serum of patients with SDMD or first-episode (FE)-SDMD. Hence, to compare the serum lipid fractions of patients with SDMD and FE-SDMD to those of a control group, the current investigation excluded both patients and controls with MetS. The specific hypothesis posits that SDMD and FE-SDMD are associated with reduced LCAT activity because Maes et al. [27] suggested that lowered LCAT could be a trait feature. Conversely, drawing from the findings of another investigation [35], we postulate that SDMD and FE-SDMD do not exhibit conspicuous abnormalities in atherogenicity profiles.

## Subjects and methods

### Participants

All the participants were enrolled academic students at Chulalongkorn University in Bangkok, Thailand, where they were enlisted in various academic departments. Recruiting occurred between November 2021 and February 2023. We recruited 108 Thai-speaking university students, of which sixty-four were depressed and forty-four were healthy control students. They were matched according to body mass index (BMI), years of education, age (within the age range of 18 to 35 years), and gender. In accordance with the inclusion and exclusion criteria, three students were omitted from the study: two from the SDMD group and one from the control group. The depressed students satisfied the DSM-5 case definition of a major depressive episode [2] requirements for MDD. Additionally, they demonstrated a Hamilton Depression Rating Scale (HAM-D) [16] score of at least 7 (excluding patients in remission) and less than 22 (to define SDMD). By applying predetermined criteria [21, 26], our study exclusively incorporated patients diagnosed with SDMD while excluding those with MDMD. Moreover, forty-seven of the 64 SDMD students included in the study exhibited a FE-SDMD.

We excluded from the study participants (both depressed and control students) who met the following criteria: a) had MetS; b) were at an increased risk of suicidal ideation; c) had medical conditions that impair immunity (e.g., autoimmune disease, inflammatory bowel disease, psoriasis, lupus erythematosus, psoriasis, rheumatoid arthritis, psoriasis, lupus erythematosus, autoimmune disease, inflammatory bowel disease, and rheumatoid arthritis; and d) had neurological disorders (including multiple sclerosis). As the study exclusively enrolled patients diagnosed with SDMD and precluded those with MDMD, there were no patients with MDD who presented with melancholia or psychotic features. Female students who were pregnant or lactating were omitted from the study. The normal control subjects who presented with a lifetime or current diagnosis of MDD, adjustment disorder with depressed mood, and dysthymia or those with a family history of MDD, bipolar disorder, and suicide in first-degree relatives were precluded from the study.

The study protocol was granted approval by the Institutional Review Board (IRB) No. 351/63 of Chulalongkorn University’s Faculty of Medicine in Bangkok, Thailand. Written informed assent was obtained from all participants prior to the commencement of the study.

### Design and Clinical measurements

Participants in this case-control study included both depressed and nondepressed students. The interviews were conducted at the King Chulalongkorn Memorial Hospital in Bangkok, Thailand, Department of Psychiatry. A research assistant (AV) who possesses extensive clinical psychology training administered the semi-structured interviews (AV). The interview incorporated socio-demographic information, a record of previous COVID-19 infections, and the frequency of depressive episodes. The severity of depression was assessed utilizing two distinct rating scales. First, the HAM-D interview [16] was translated in Thai and validated [25]. Second, we employed the Beck Depression Inventory-II (BDI-II) as a self-rating scale to assess the severity of depression [6]. Mungpanich [38] translated and validated the BDI-II in Thai.

The assessment of suicidal behaviors (SBs) was conducted through the utilization of a semi-structured interview incorporating the Columbia-Suicide Severity Rating Scale (C-SSRS) [46] in a validated Thai translation [47]. We utilized the C-SSRS to evaluate both current (within one month) and past (up to one month prior to participant inclusion) SBs, which are classified as suicidal ideation (SI) and attempts (SA). As previously described, we calculated lifetime SB scores by combining past and present SI and SA scores derived from C-SSRS ratings [63]. By extracting the first validated principal component from the HAM-D, BDI-II, and SB scores, the depression phenome score was calculated [63]. The severity of rumination, with a specific focus on brooding, was evaluated using the Ruminative Response Scale (RRS) [40]. A Thai-validated translation of the RRS was utilized [61]. In a prior investigation, we established that the most critical element of rumination was brooding; thus, in the present study, we exclusively employed the brooding score [21]. Because brooding contributes to a portion of the variability in the depression phenome, we additionally investigated the impact of lipid biomarkers on the phenome after subtracting the effects of brooding (using regression analysis). Consequently, the impacts of biomarkers on the residualized phenome scores, which were modified to account for brooding, were calculated.

MetS was defined by the 2009 Joint Scientific Statement of the American Heart Association and the National Heart, Lung, and Blood Institute (43) as the concurrent presence of at least three of the subsequent components: (1) A waist circumference of at least 90 cm for men and 80 cm for women, or a body mass index (BMI) of at least 25 kg/m2; (2) A high triglyceride level of at least 150 mg/dL; (3) A low HDL cholesterol level of at least 40 mg/dL for men and 50 mg/dL for women; (4) A high blood pressure of at least 130 mm Hg for systolic blood pressure and ≥ 85 mm Hg for diastolic blood pressure; (5) A fasting blood glucose level of at least 100 mg/dL or a diabetes diagnosis. The body mass index (BMI) is computed by dividing weight in kilograms by height in meters squared. We examined the effects of “subclinical MetS” using two variables: a) an ordinal score ranging from no criteria of subclinical MetS to one criterion (representing stage 1 subclinical MetS) to two criteria (stage 2 subclinical MetS); and b) a binary variable distinguishing between stages zero versus 1+2.

### Biomarker measurements

A cumulative of thirty milliliters of fasting venous blood were obtained from each student using a disposable syringe and serum tubing during the hours of 8:00 to 11:00 a.m. Serum was extracted after blood centrifugation at 35,000 revolutions per minute. Small aliquots were placed in Eppendorf receptacles and stored at −80 degrees Celsius until they were thawed prior to biomarker assays. The total cholesterol, HDLc, triglyceride, and direct LDLc concentrations were determined utilizing the Alinity C (Abbott Laboratories, USA; Otawara-Shi, Tochigi-Ken, Japan) as previously documented [21, 35]. The respective coefficients of variation for total cholesterol (TC), HDLc, LDLc, and triglycerides were 2.3%, 2.6%, 2.3%, and 4.5%, respectively. The concentrations of ApoA1 and ApoB were ascertained via immunoturbidimetric assays employing the Roche Cobas 6000 and c501 module (Roche, Rotkreuz, Switzerland). The intra-assay coefficients of variation values for Apo A1 and Apo B were 1.75% and 2.64%, respectively. The quantity of free cholesterol (FC) and ApoE were determined utilizing the Free Cholesterol Colorimetric Assay Kit (Elabscience, cat number: E-BC-K004-M) and the Human ApoE (Apolipoprotein E) Elisa kit, respectively. The intra-assay coefficients of variation were 1.9% and 4.67%, respectively.

The formula utilized to calculate the esterified cholesterol ratio, an indicator of LCAT activity, was (1 - free cholesterol / total cholesterol) x 100 (1 Maes et al., 1994). The ApoB/ApoA ratio was calculated as a measure of atherogenicity. Maes et al. (27=2023) calculated an additional atherogenicity index in the form of a z unit-based composite score: z transformation of z ApoB + z triglycerides + z free cholesterol + z LDLc. Additionally, an RCT index was calculated as z HDLc + z LCAT + z ApoA1. Two indices of atherogenicity and anti-atherogenicity were calculated as follows: a) z triglycerides + z ApoB + z free cholesterol + z LDLc - z HDLc - z LCAT - z ApoA1 (Athero/RCT); or z (z triglycerides + z ApoB + z free cholesterol + z LDLc) – z LCAT (Athero/LCAT).

### Statistical analyses

Utilizing Pearson’s product-moment correlation coefficients or Spearman’s rank order correlation coefficients, the analysis investigated the correlations between continuous variables. To evaluate the statistical relationships between categorical variables, a contingency table analysis (X^2^-test) was employed. Analysis of variance (ANOVA) was utilized by the researchers to investigate the associations between diagnostic categories and clinical data. The study utilized univariate and multivariate generalized linear models (GLMs) to examine the associations between lipids and lipid composite scores and SDMD or FE-SDMD. The researchers utilized both manual and automatic multivariable regression analysis to identify the most significant biomarkers (input data) of the output data (the clinical phenome data). An automated regression analysis was performed, utilizing p-to-entry and p-to-remove thresholds of 0.05 and 0.06, respectively. Various model metrics were computed, including F, df, p-values, and variance explained (R^2^, which served as the model’s effect size). Additionally, parameter estimates for significant predictors were derived, including standardized beta coefficients accompanied by t and p values. Furthermore, an examination of the data was conducted to identify any collinearity or multicollinearity concerns by employing variance inflation factors (VIF), tolerance, condition indices, and variance proportions. The homoskedasticity was assessed by employing the White and modified Breusch-Pagan tests. To examine the distribution of the residuals from the regression, standardized residual plots were displayed. Adjusting for other confounding predictors, we produced partial regression diagrams that illustrate the result of partial regression of the dependent variable on a single predictor. Statistical significance was ascertained by employing two-tailed tests and a p-value of 0.05. Version 29 of SPSS for Windows was utilized to analyze the data.

By employing SmartPLS, Partial Least Squares (PLS) analysis was utilized [50] to investigate the causal connections between the lipid data, brooding, and the manifestation of depression. A total of 5,000 bootstrap samples were utilized to conduct complete PLS analysis. This analysis was restricted to models whose inner and outer models met predetermined quality criteria: a) the Standardized Root Mean Square Residual (SRMR) of the model should fall below 0.08; b) the extracted factors must exhibit satisfactory composite and convergence reliability as measured by Cronbach’s alpha (> 0.7), rho A values (> 0.75), and average variance extracted (> 0.5); c) all loadings on the extracted factors should surpass 0.7; d) Confirmatory Tetrad Analysis must show that the factors are not mis-specified as reflective models; and e) all variables should display discriminant validity as assessed with the Heterotrait-Monotrait ratio (HTMT). Complete PLS analysis is then conducted, and path coefficients (accompanied by p-values), total effects, total indirect effects, and specific indirect effects are computed.

Multiple regression analysis and PLS analysis were utilized as the primary data analyses in this research to investigate the correlations between the depression phenome score (output variables) and the biomarkers (input variables). Because of the sample size (n=108), only five input variables were permitted. The determination of the minimal required sample size (n=79) is based on several factors: an effect size of 0.176, which explains approximately 15% of the variance, a desired statistical power of 0.08, the incorporation of 5 covariates, and a significance level of 0.05 (G*Power 3.1.9.7).

## Results

### Sociodemographic and clinical data

**Table 1** shows the sociodemographic and clinical data of the 108 students that were recruited to participate in the current study. There were no significant differences in age, gender, BMI, years of education, marital status, smoking or alcohol use, or a prior diagnosis of mild COVID-19 between students with SDMD and normal control students. The total HAM-D, total BDI-II, SB, phenome, and brooding scores were significantly higher in SDMD students than in controls. The residualized phenome data obtained by regression on brooding were significantly higher in SDMD patients than in controls.

**Table 1.** Sociodemographic, clinical, and metabolic data of students with simple dysmood disorder (SDMD) and control students (HC).

| **Biomarkers** | **HC (n=44)** | **SDMD (n=64)** | **F/X<sup>2</sup>** | **df** | **p** |
| --- | --- | --- | --- | --- | --- |
| Age (years) | 23.5 (3.2) | 22.4 (3.2) | 2.75 | 1/106 | 0.100 |
| Sex (F/M) | 37/7 | 54/10 | 0.00 | 1 | 0.968 |
| Education (years) | 16.6 (1.5) | 15.9 (2.7) | 2.15 | 1/106 | 0.146 |
| BMI (kg/m<sup>2</sup>) | 22.3 (3.6) | 22.4 (5.1) | 0.01 | 1/106 | 0.927 |
| Single/relationship or marriage | 27/17 | 27/37 | 3.84 | 1 | 0.050 |
| Smoking (No/Yes) | 43/1 | 59/5 | 1.53 | 1 | 0.217 |
| Student drinking (No/Yes) | 41/3 | 53/11 | 2.49 | 1 | 0.115 |
| Suffered from mild COVID-19 (No/Yes) | 30/14 | 49/15 | 0.93 | 1 | 0.334 |
| Total HAM-D score | 2.4 (2.0) | 14.6 (4.7) | 257.60 | 1/106 | <0.001 |
| Total BDI-II score | 6.7 (5.4) | 24.7 (11.5) | 86.85 | 1/106 | <0.001 |
| SB score (z score) | -0.872 (0.199) | 0.599 (0.881) | 118.13 | 1/106 | <0.001 |
| Phenome (z score) | -0.998 (0.300) | 0.686 (0.682) | 236.91 | 1/106 | <0.001 |
| Brooding (z score) | -0.802 (0.809) | 0.551 (0.702) | 85.57 | 1/106 | <0.001 |
| Residualized phenome (z score) | -0.716 (0.091) | 0.482 (0.071) | 85.82 | 1/105 | <0.005 |

All data are shown as mean (SD) or as ratios. F: results of analysis of variance; X<sup>2</sup>: results of contingency analysis.
HAM-D: Hamilton Depression Rating Scale; BDI-II: Beck Depression Inventory; SB: suicidal behaviors; Residualized phenome: brooding subtracted phenome scores (following regression analysis)

### Lipids in SDMD and FE-SDMD

**Table 2** shows the lipid measurements in both patients and controls. ESF, Table 1 shows the raw scores of the lipid measurements in both study groups (after covarying for age, sex, and BMI). Using univariate GLM analysis (covaried for age, sex, and BMI), we found that free cortisol and ApoE were significantly higher in SDMD than in controls, whilst the LCAT index was significantly decreased in patients. The atherogenicity composite was significantly higher in SDMD patients as compared with controls as were the Athero/LCAT and Athero/RCT indices.

**Table 2.** Assessment of the lipid variables in students with simple dysmood disorder (SDMD) and healthy control students (HC).

| Variables (z scores) | HC | SDMD | F (df=1/98) | p |
| --- | --- | --- | --- | --- |
| Total cholesterol | -0.184 (0.150) | 0.109 (0.125) | 2.24 | 0.139 |
| HDLc | -0.052 (0.133) | 0.072 (0.111) | 0.502 | 0.480 |
| Triglycerides | -0.127 (0.145) | 0.133 (0.121) | 1.58 | 0.212 |
| LDLc | -0.152 (0.152) | 0.111 (0.128) | 1.73 | 0.192 |
| Free cholesterol | -0.500 (0.133) | 0.352 (0.111) | 23.86 | <0.001 |
| Apo A | -0.171 (0.148) | 0.129 (0.124) | 2.39 | 0.126 |
| Apo B | -0.178 (0.140) | 0.097 0.117 | 2.22 | 0.139 |
| ApoB/ApoA | -0.106 (0.129) | 0.039 0.108 | 0.72 | 0.398 |
| Apo E | -0.293 (0.122) | 0.172 0.102 | 8.39 | 0.005 |
| LCAT index | 0.435 (0.138) | -0.241 0.116 | 13.85 | <0.001 |
| RCT index | 0.102 (0.138) | -0.017 0.116 | 0.43 | 0.512 |
| Atherogenicity index | -0.366 (0.134) | 0.265 0.122 | 11.59 | 0.001 |
| Athero/LCAT ratio | -0.472 (0.124) | 0.388 0.104 | 27.96 | <0.001 |
| Athero/RCT ratio | -0.306 (0.124) | 0.216 0.104 | 10.24 | 0.002 |

Results are shown as mean (SE). All results of univariate GLM analyses with age, sex, and body mass index as covariates.
HDLc: high density lipoprotein cholesterol; LDLc: low density lipoprotein cholesterol; Apo: apolipoprotein; LCAT: lecithin-cholesterol acyltransferase; RCT: reverse cholesterol transport; Athero: atherogenicity index.

Some patients were treated with venlafaxine (n=7), sertraline (n=20), escitalopram (n=16), fluoxetine (n=10), atypical antipsychotics (n=12), or benzodiazepines (n=29). Therefore, we have statistically adjusted for the drug state of the patients to exclude any effects of the drug state on the results. We could not find any significant effects of venlafaxine (p=0.083), sertraline (p=0.614), escitalopram (p=0.465), fluoxetine (p=0.298), atypical antipsychotics (p=0.472), and benzodiazepines (p=0.954). Covarying for the use of these psychotropic drugs did not change the association between the lipid data and the phenome data. Only six subjects occasionally used NSAIDS and six others paracetamol. Both drugs had no significant effects on any of the lipid concentrations (tested at a p=0.05 level, without p correction) and covarying for these drugs showed no significant change in any of the results shown in Table 2.

**Table 3** shows the same variable but now in FE-SDMD patients versus controls. The results show the same pattern as with the total SDMD group.

**Table 3.** Assessment of the lipid variables in students with first episode (FE) simple dysmood disorder (SDMD) and healthy control students (HC).

| Variables (z scores) | HC (n=43) | FE-SDMD (n=45) | F (df=1/82) | p |
| --- | --- | --- | --- | --- |
| Total cholesterol | -0.189 0.154 | 0.081 0.151 | 1.52 | 0.221 |
| HDLc | -0.023 0.129 | 0.119 0.126 | 0.61 | 0.438 |
| Triglycerides | -0.167 0.141 | 0.275 0.138 | 4.86 | 0.030 |
| LDLc | -0.158 0.152 | 0.006 0.148 | 0.58 | 0.450 |
| Free cholesterol | -0.505 0.129 | 0.333 0.126 | 21.03 | <0.001 |
| ApoA1 | -0.153 0.151 | 0.123 0.148 | 1.66 | 0.201 |
| ApoB | -0.183 0.139 | -0.082 0.136 | 0.26 | 0.611 |
| ApoB/ApoA ratio | -0.120 0.121 | -0.104 0.118 | 0.01 | 0.926 |
| ApoE | -0.326 0.119 | 0.163 0.116 | 8.43 | 0.005 |
| LCAT index | 0.432 0.141 | -0.281 0.138 | 12.64 | <0.001 |
| RCT index | 0.122 0.140 | -0.016 0.137 | 0.49 | 0.487 |
| Atherogenicity index | -0.361 0.133 | 0.222 0.130 | 9.52 | 0.003 |
| Athero/LCAT ratio | -0.488 0.118 | 0.351 0.115 | 25.04 | <0.001 |
| Athero/RCT ratio | -0.339 0.113 | 0.184 0.111 | 10.53 | 0.002 |

Results are shown as mean (SE). All results of univariate GLM analyses with age, sex, and body mass index as covariates.
HDLc: high density lipoprotein cholesterol; LDLc: low density lipoprotein cholesterol; Apo: apolipoprotein; LCAT: lecithin-cholesterol acyltransferase; RCT: reverse cholesterol transport; Athero: atherogenicity index.

### SDMD and subclinical MetS

ESF, Table 2 shows the lipids data in stage 0, stage 1, and stage 2 subclinical MetS. HDLc was significantly lower in those with the stage 1 and 2 subclinical MetS as compared with stage 0. Triglyceride levels were higher in stage 2 than in stage 1 and stage 0 subclinical MetS. ApoE was significantly different between the three stages and increased from stage 0 to stage 1 to stage 2. The ApoB/ApoA ratio was significantly higher in stage 2 than stage 1 and stage 0. RCT was lower in both stages 1 and 2 compared to stage 0. The Athero/RCT index was significantly higher in stage 2 than in the other two stages.

ESF, Table 3 shows the interactions between the diagnosis SDMD and the three stages of subclinical MetS. This interaction pattern was significant for HDLc, ApoA, ApoB/ApoA1 ratio, RCT and Athero/RCT. **Figure 1** and ESF, Table 3 show that no such interaction was established for free cholesterol, which was consistently higher in SDMD than in controls all over the three subclinical MetS stages. In subclinical MetS stage 0, free cholesterol was significantly higher in SDMD (F=15.75, df=1/63, p<0.001) and FE-SDMD (F=16.42, df=1/55, p<0.001) than in controls. An opposite pattern was established for LCAT, which was consistently lowered in all three subclinical MetS stages of SDMD than controls (**Figure 2**). In subclinical MetS stage 0, LCAT was significantly lower in SDMD (F=4.53, df=1/63, p=0.037) and FE-SDMD (F=3.68, df=1/55, p=0.06, one tailed test) than in controls. ESF, Figures 1–5 show the significant interaction patterns between SDMD and subclinical MetS. SDMD patients with subclinical MetS stage 2 show significant decreases in HDLc, ApoA1 and RCT and increases in the ApoB/ApoA1 and Athero/RCT ratios.

**Figure 1.**
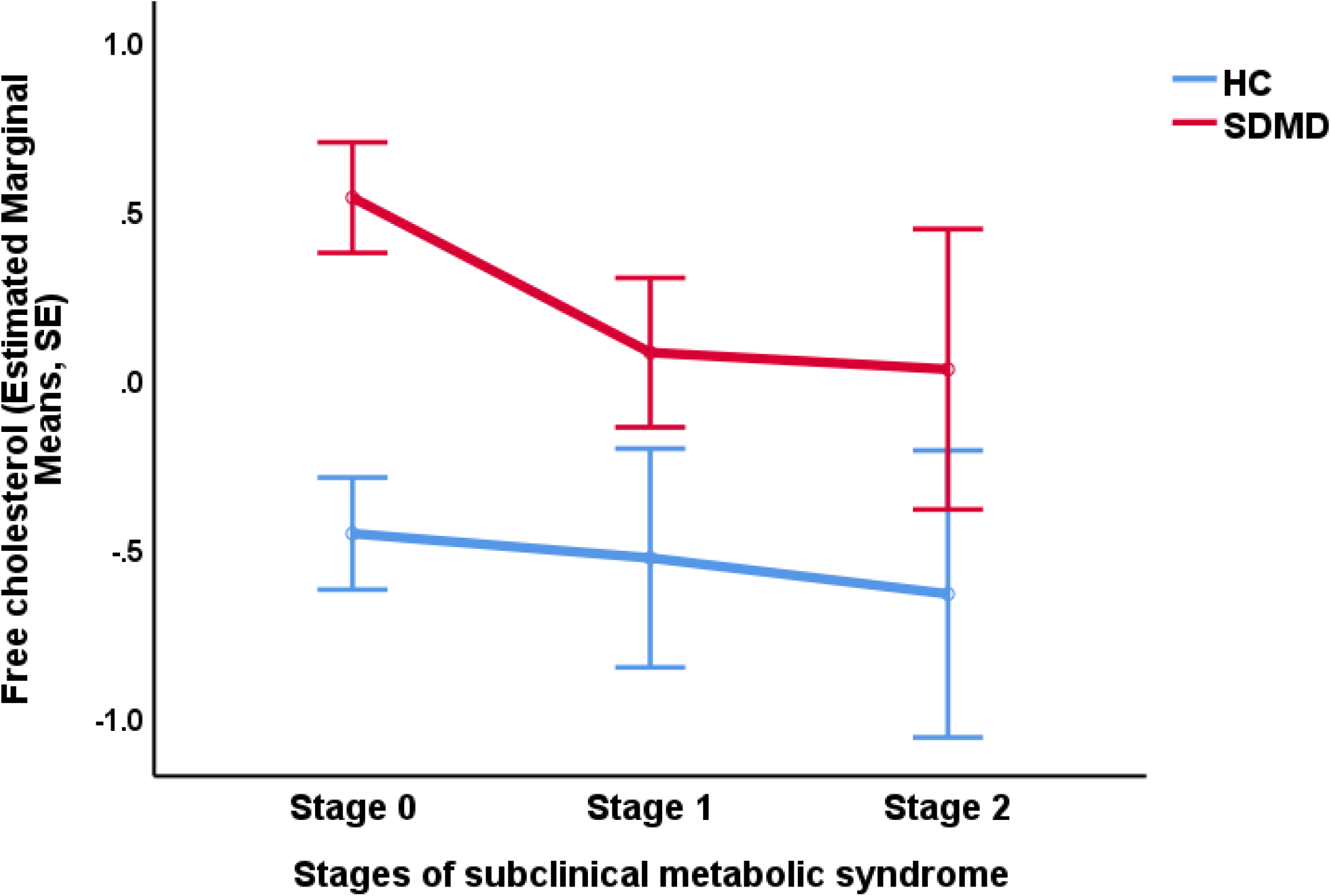
Serum free cholesterol levels in students with simple dysmood disorder (SDMD) and healthy control students (HC) in the three stages of subclinical metabolic syndrome. These levels were significantly higher (p<0.001) in SDMD than in controls in stage 0 of the subclinical metabolic syndrome.

**Figure 2.**
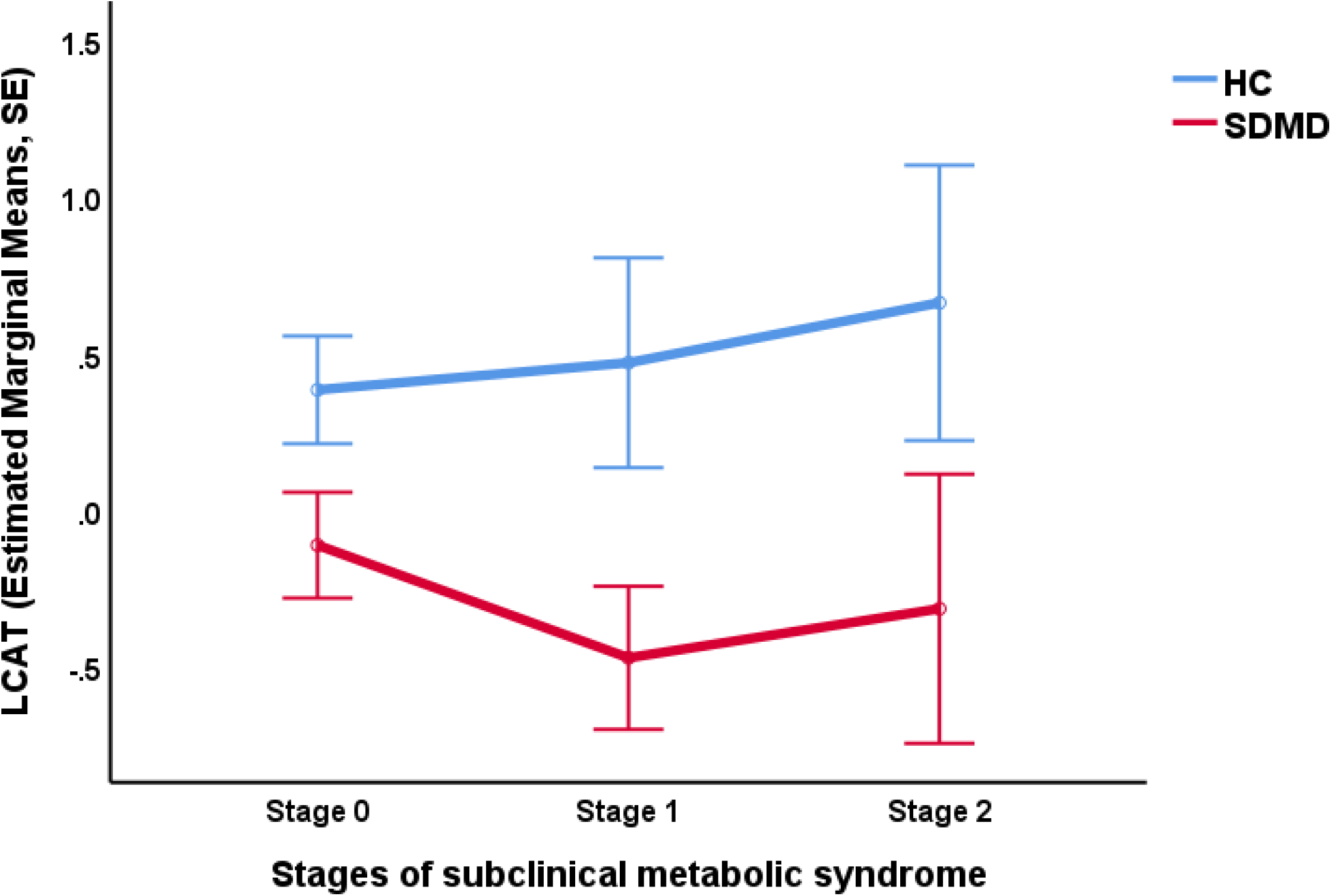
Serum lecithin-cholesterol acyltransferase (LCAT) levels in students with simple dysmood disorder (SDMD) and healthy control students (HC) in the three stages of subclinical metabolic syndrome. These levels were significantly higher (p=0.037) in SDMD than in controls in stage 0 of the subclinical metabolic syndrome.

These results show that one should adjust even for those stages on subclinical MetS when examining lipid profiles. Likewise, we used subclinical MetS as a binary variable or three group variable (the three stages) in Tables 2 and 3 and detected that BMI was a more significant predictor of lipids and that after considering the effects on BMI the effects of subclinical MetS disappeared.

### Correlations between lipid variables and clinical phenome data

**Table 4** shows the correlation matrix among clinical rating scale scores and the key lipid variables. The HAM-D, BDI, brooding, SBs and phenome scores were significantly associated with free cholesterol concentrations (positively) and the LCAT index (inversely). **Figure 3** shows the partial regression of the phenome on free cholesterol (after adjusting for age, sex, and BMI). The RCT index was significantly and inversely associated with brooding. The atherogenicity index was significantly associated with the HAM-D, BDI, and phenome scores. The Athero/LCAT and Athero/RCT ratios were significantly correlated with all five clinical variables. **Figure 4** shows the partial regression of the phenome on the Athero/LCAT ratio (after adjusting for age, sex, and BMI).

**Figure 3.**
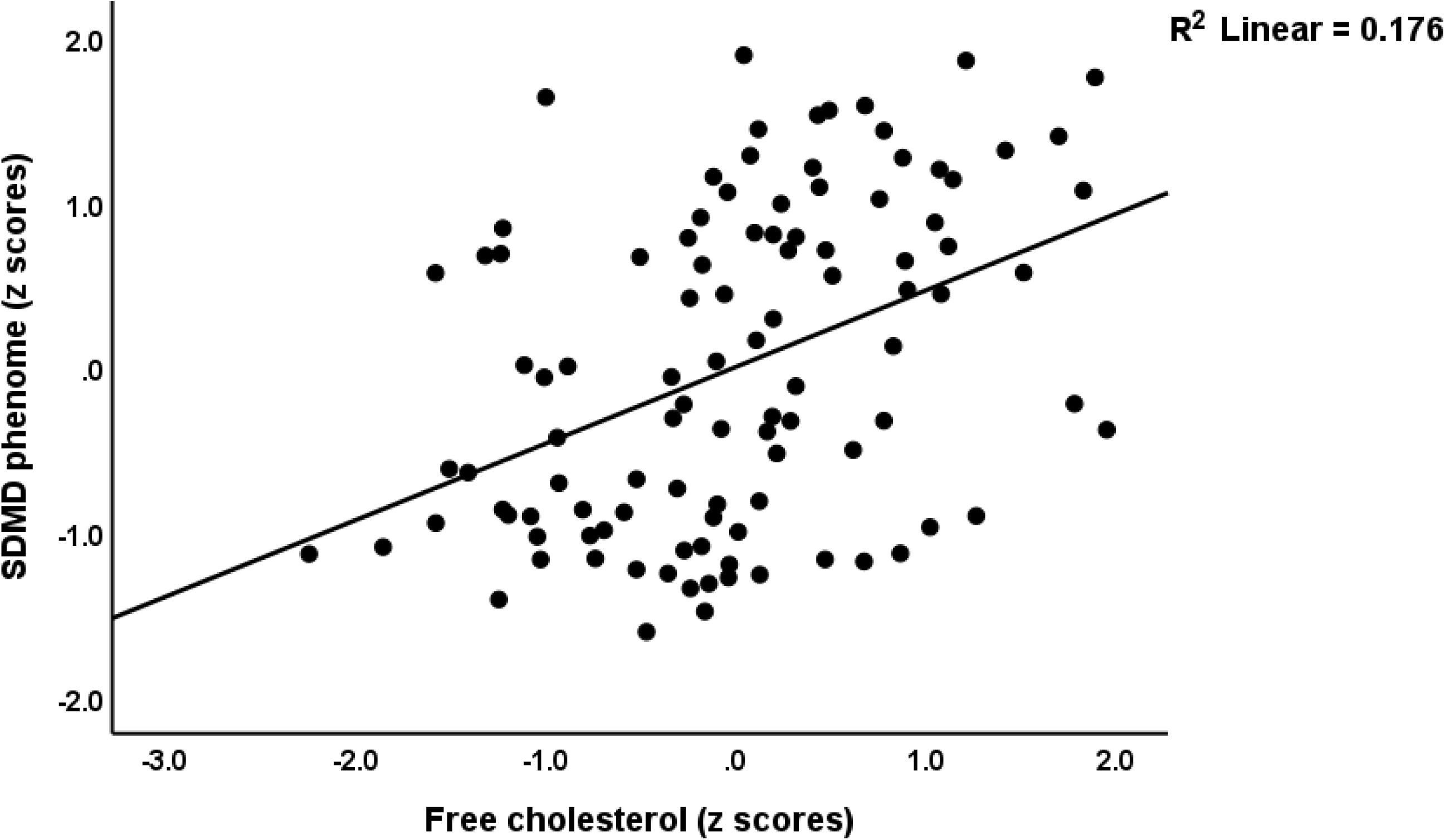
Partial regression of the phenome of simple dysmood disorder (SDMD) on free cholesterol (after adjusting for age, sex, and body mass index).

**Figure 4.**
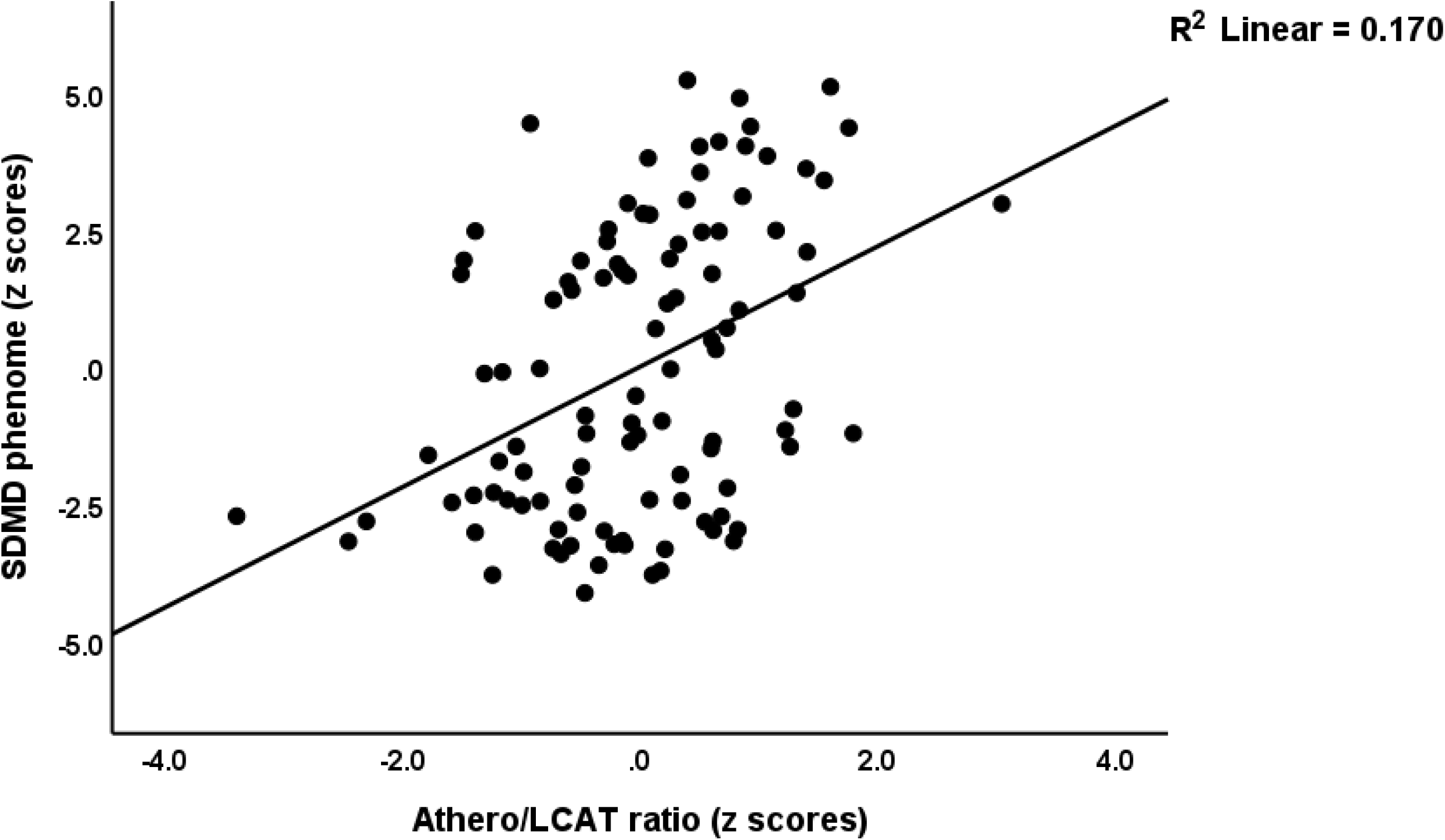
Partial regression of the phenome of simple dysmood disorder (SDMD) on the atherogenicity (Athero) / lecithin-cholesterol acyltransferase (LCAT) ratio (after adjusting for age, sex, and body mass index).

**Table 4.** Correlation matrix among clinical rating scale scores and lipid variables.

| **Variables** | **HAM-D** | **BDI-II** | **Brooding** | **SB** | **Phenom** |
| --- | --- | --- | --- | --- | --- |
| Free cholesterol | **0.383 (<0.001)** | **0.313 (0.001)** | **0.255 (0.008)** | **0.283 (0.003)** | **0.360 (<0.001)** |
| LCAT index | **-0.388 (<0.001)** | **-0.297 (0.002)** | **-0.301 (0.002)** | **-0.283 (0.003)** | **-0.358 (<0.001)** |
| RCT index | -0.153 (0.118) | -0.136 (0.164) | **-0.238 (0.014)** | -0.187 (0.055) | -0.176 (0.071) |
| Atherogenicity index | **0.227 (0.020)** | **0.200 (0.041)** | 0.124 (0.208) | 0.183 (0.062) | **0.225 (0.006)** |
| Athero/LCAT index | **0.407 (<0.001)** | **0.325 (0.001)** | **0.279 (0.004)** | **0.323 (0.001)** | **0.389 (<0.001)** |
| Athero/RCT index | **0.251 (0.010)** | **0.215 (0.028)** | **0.219 (0.025)** | **0.253 (0.010)** | **0.266 (0.006)** |

Shown are Pearson's product moment correlation coefficients (all n=108).
LCAT: lecithin-cholesterol acyltransferase; RCT: reverse cholesterol transport; Athero: atherogenicity.

ApoE was significantly correlated with free cholesterol (r=0.294, p=0.002), total cholesterol (r=0.014), triglycerides (r=0.514, p<0.001), ApoB (r=0.354, p<0.001) and HDLc (r=−0.198, p=0.041). Free cholesterol was significantly correlated with total cholesterol (r=0.645, p<0.001), triglycerides (r=0.254, p=0.009), LDLc (r=0.546, p<0.001), ApoB (r=0.500, p<0.001)

### Multiple regression analysis

**Table 5** shows the results of multiple regression analyses with the clinical phenome scores as output variables and lipid biomarkers as input variables. We show types of results for each dependent variable, namely one without inclusion of brooding as explanatory variable (the “a” regressions), and one with inclusion of brooding (the “b” regressions). Table 5, regression 1a shows that 19.7% of the variance in the HAM-D score was explained by free cholesterol and the LCAT index. After allowing for the effects of grooming, we found that both grooming and free cholesterol predicted the HAM-D score (regression 1b). Regression 2a shows that the BDI-II score was predicted (17.1%) by free cholesterol (positively) and HDLc and education (inversely. Entering brooding in the same analysis showed that ApoB is significantly associated with the BDI score. Regression #3a shows that 16.2% of the variance in SBs is explained by the Athero/LCAT ratio and education, whilst regression #3b shows that brooding coupled with the same ratio and education explain 41.4% of the variance. As with the BDI score, the best predictors of the phenome score were free cholesterol, education, and HDL (regression #4a). Regression #4b shows that 63.3% of the variance in the phenome is explained by brooding, free cholesterol and age combined. We found that 7.1% of the variance in brooding was explained by LCAT.

**Table 5.**
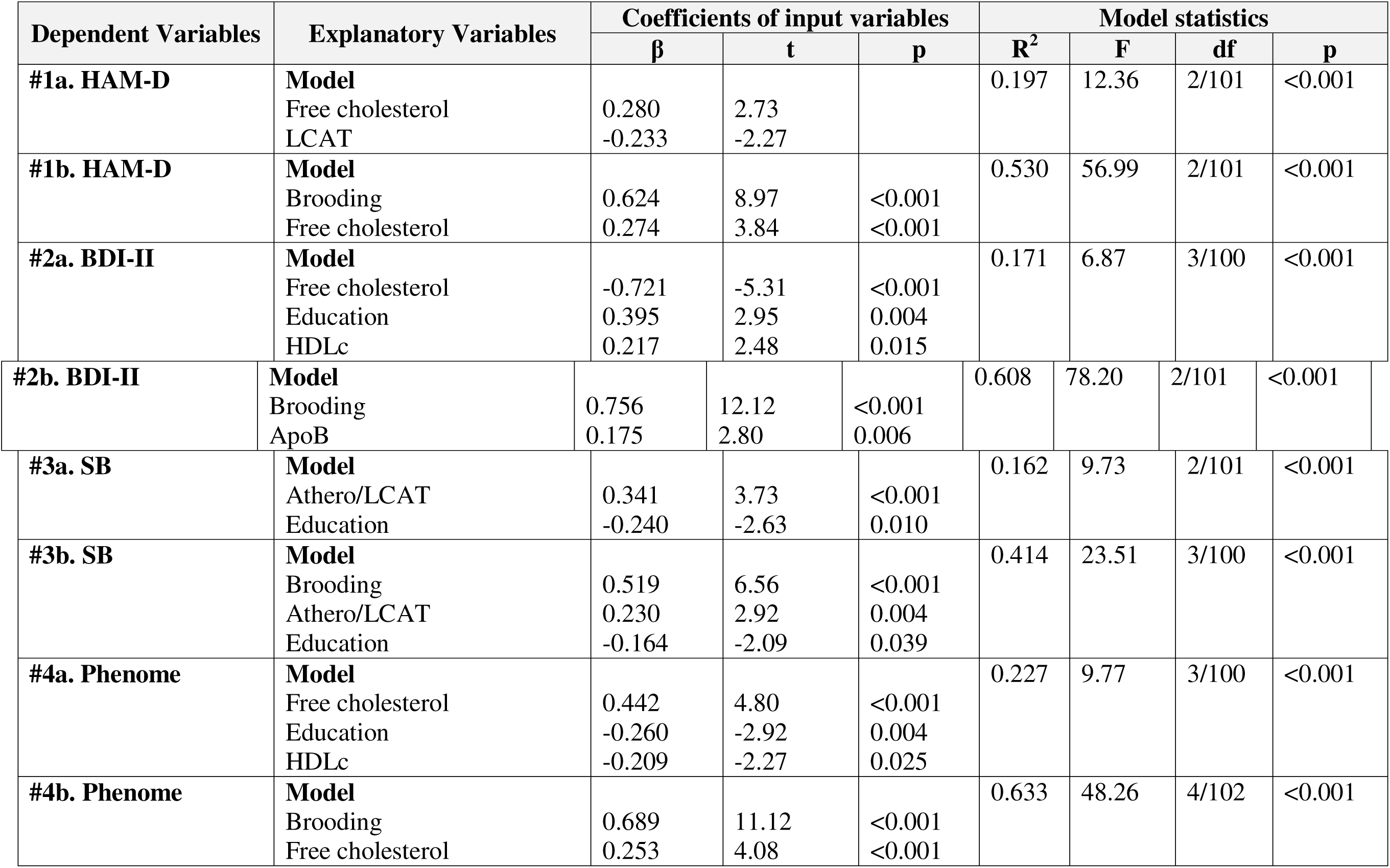
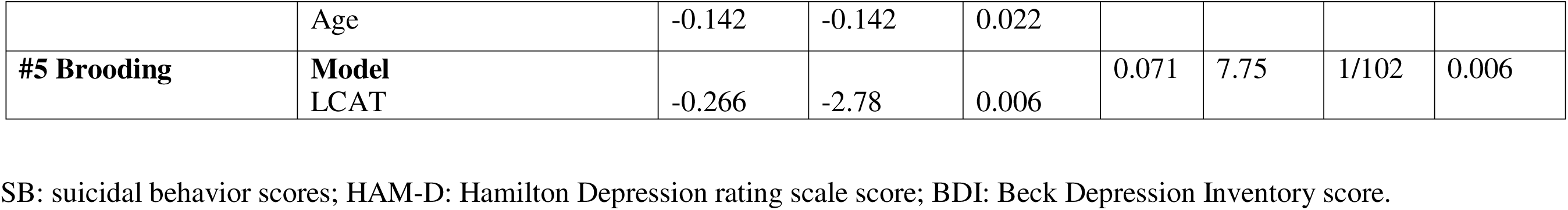
Results of multiple regression analyses with suicidal behaviors or clinical scores as dependent variables and immune biomarkers as explanatory variables.

### Results of PLS analysis

To decipher whether LCAT, free cholesterol or APOE are best associated with the phenome of depression, we conducted a PLS analysis with the phenome of depression as final outcome and with brooding, free cholesterol, LCAT, and ApoE as direct explanatory variables. In addition, LCAT was allowed to predict free cholesterol and brooding, which thus were allowed to function as mediators. The phenome was entered as a factor extracted from HAM-D, BDI and SBs scores. **Figure 5** shows the final PLS model which shows the significant paths only. Sex, age, and subclinical metabolic syndrome stages were allowed to predict all other variables. The model displayed in Figure 5 shows adequate model fit criteria including SRMR of 0.025, whilst also the extracted factor is more than adequate with AVE=0.806, Cronbach’s alpha=0.880, and composite reliability=0.891. We found that 62.2% of the variance in the phenome was predicted by free cholesterol, brooding and age; 12.0% of the variance in brooding was explained by ApoE and LCAT; 25.3% of the variance in free cholesterol by age, and LCAT; 28.9% of the variance in ApoE by sex and subclinical MetS; and 3.2% of the variance in LCAT by age. This analysis shows specific indirect effects of LCAT of the phenome mediated via brooding (t=−3.17, p=0.001) and free cholesterol (t=−2.85, p=0.002). There were no direct effects of LCAT on the phenome, but the total effect of LCAT was highly significant (t=−4.21, p<0.001). The specific indirect effects of ApoE on the phenome were mediated by brooding (t=1.92, p=0.027). Subclinical MetS did not have a significant effect on the phenome (t=1.64, p=0.051).

**Figure 5.**
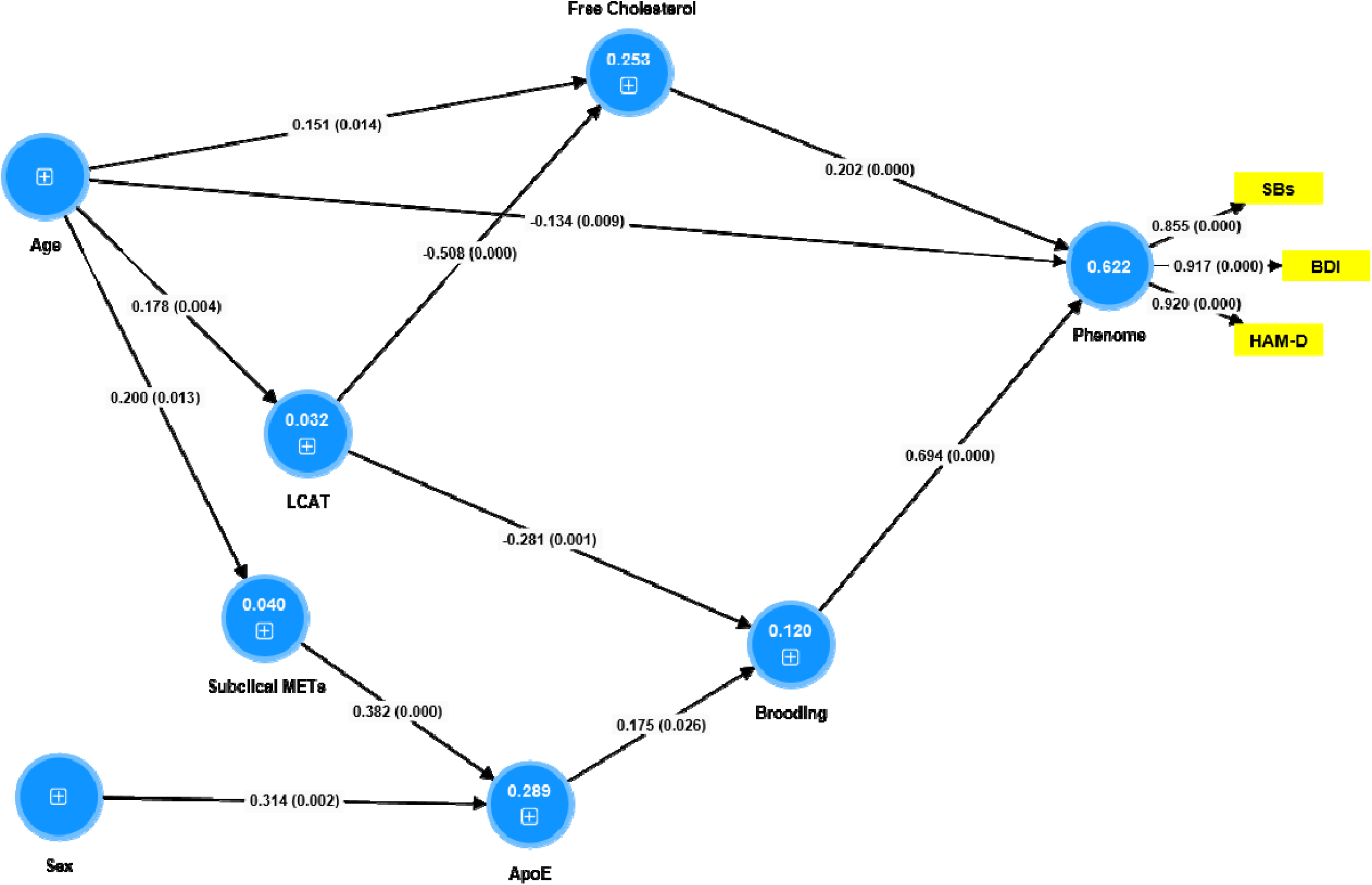
Partial Least Squares (PLS) model which shows the significant paths from lipid variables to the phenome of depression. The latter is conceptualized as a factor extracted from the total scores on the Hamilton Depression Rating (HAM-D) and Beck Depression Inventory (BDI) scores and suicidal behaviors (SBs). We allowed all indicators to predict the phenome, lecithin-cholesterol acyltransferase (LCAT) to predict free cholesterol, all lipid variables to predict brooding, and age, sex, and subclinical metabolic syndrome to predict all other indicators. This model only displays the significant paths and indicators. Shown are path coefficients with exact p values and the loadings with exact p values. White figures in blue circles denote the explained variance.

To examine the effects of LCAT on the phenome in a later stage of SDMD and MDMD, we have re-analyzed the data of Maes et al. (2023). Figure 6 shows this PLS model which examined the direct and indirect effects of LCAT on the phenome. We found significant indirect effects of LCAT on the phenome that were completely mediated via increasing atherogenicity (t=−0.24, p=0.019). There were no direct effects of LCAT on the phenome (p=0.379).

**Figure 6.**
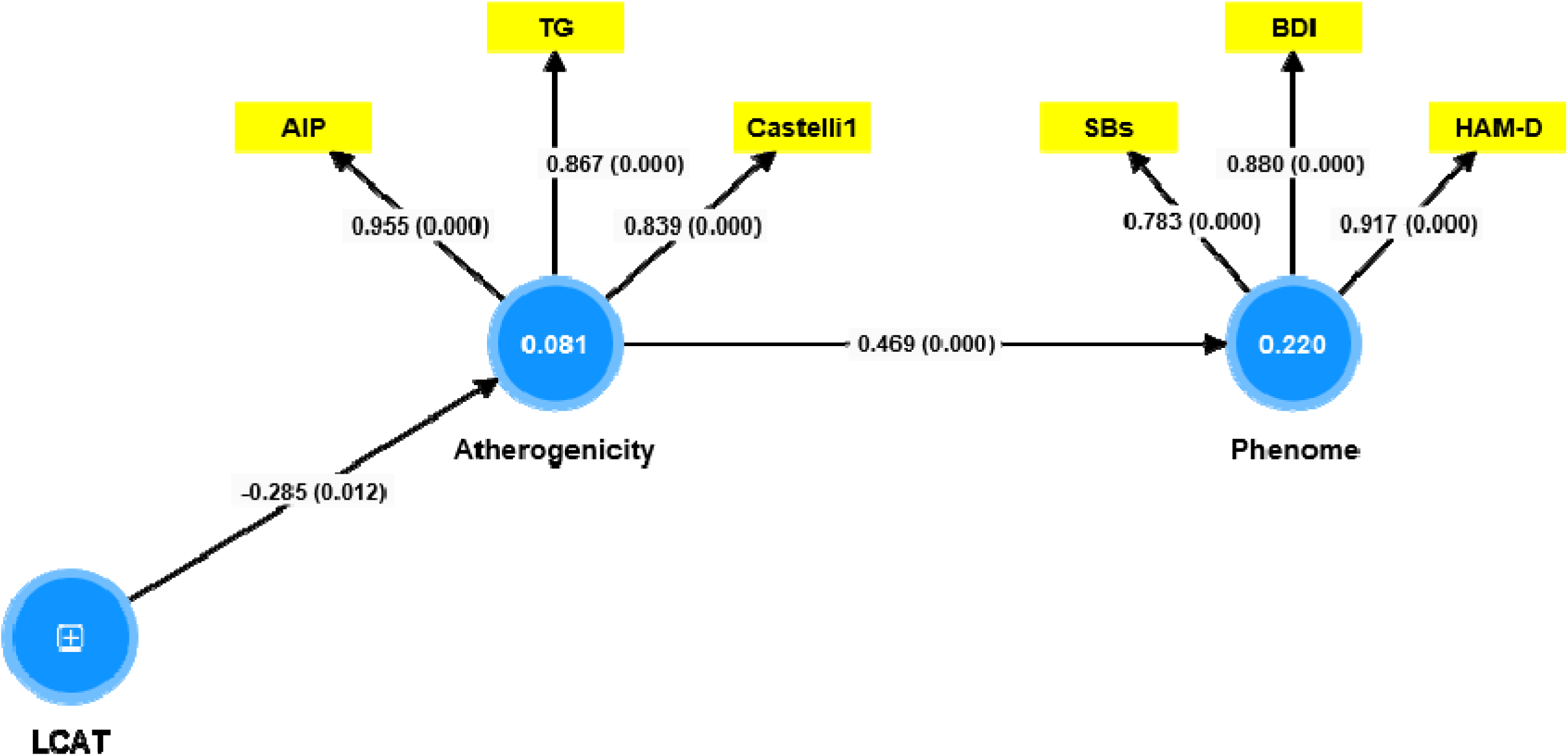
Partial Least Squares (PLS) model which shows the significant paths from lipid variables to the phenome of depression. The latter is conceptualized as a factor extracted from the total scores on the Hamilton Depression Rating (HAM-D) and Beck Depression Inventory (BDI) scores and suicidal behaviors (SBs). Atherogenicity was conceptualized as a factor extracted from plasma triglyceride levels, the Castelli 1 index (total cholesterol / high-density lipoprotein cholesterol or HDLc), and the atherogenic index of plasma (AIP; triglycerides/HDLc). Lecithin-cholesterol acyltransferase (LCAT) was allowed to predict both factors. This model only shows the significant paths and indicators. Shown are path coefficients with exact p values and the loadings with exact p values. White figures in blue circles denote the explained variance. The data set used for this PLS analysis was presented in Maes M, Jirakran K, Vasupanrajit A, Boonchaya-Anant P, Tunvirachaisakul C. Towards a major methodological shift in depression research by assessing continuous scores of recurrence of illness, lifetime and current suicidal behaviors and phenome features: focus on atherogenicity and adverse childhood experiences medRxiv 2023.02.26.23286462; doi: https://doi.org/10.1101/2023.02.26.23286462 (This study was approved by the Chulalongkorn University Medical Faculty’s Institutional Review Board (#445/63) in Bangkok, Thailand).

## Discussion

### Lipids in FE-SDMD and SDMD

The first major finding of this study is that LCAT was decreased, and free cholesterol and ApoE increased in subjects with SDMD and FE-SDMD as compared with healthy controls. In SDMD, we could not detect significant changes in HDLc, ApoA1, RCT, ApoB and triglycerides. A previous investigation documented a tendency for atherogenicity to increase and RCT to decrease in SDMD as compared to the control group [35]. In the latter study, patients diagnosed with MDMD exhibited elevated levels of free cholesterol, triglycerides, LDLc, ApoB, and atherogenity indices, while HDLc and RCT were significantly reduced [35]. However, it is crucial to acknowledge the variations in the study samples utilized in both research studies to fully comprehend the pathogenesis of depression. In the current study, most patients were experiencing their initial episode of depression and displayed a maximum of two episodes. In contrast, our previous study group [35] comprised numerous patients with a high ROI index, indicating multiple episodes and suicidal behaviors. This is significant because an increase in ROI is associated with a reduction in RCT and an increase in atherogenicity [35]. Second, the current study exclusively enrolled patients who were in an acute phase of very mild MDD (SDMD), whereas the preceding article included patients with both SDMD and MDMD who were experiencing partial remission.

Moreover, the current study detected that in SDMD, suicidal behaviors and severity of illness were significantly associated with lowered LCAT and increased free cholesterol and derived atherogenicity indices. Previously, we discussed that lowered HDLc in the acute phase of MDD and lowered RCT and increased atherogenicity in MDMD are associated with suicidal behaviors and severity of depression [30, 35].

### SDMD and subclinical MetS

Patients with MetS were precluded from the current study, as our prior research demonstrated that the relationship between lipids and depression could only be adequately investigated in the depression subgroup devoid of MetS. Therefore, only patients without MetS were allowed to participate in both investigations, and additionally the results of both studies were adjusted for BMI. An aspect that distinguishes the present investigation is the inclusion of adjustments for subclinical MetS. It is noteworthy that the onset of subclinical MetS symptoms had a significant impact on lipid measurements. The key features of subclinical MetS were lowered HDLc, increased triglycerides and ApoE levels, whilst the LCAT index and free cholesterol remained unchanged.

Most significantly, even SDMD and FE-SDMD patients devoid of subclinical MetS exhibited the key lipid disorders of SDMD, namely elevated free cholesterol and decreased LCAT. This indicates that these aberrations in lipid metabolism are already present prior to the onset of subclinical MetS. Conversely, elevated levels of ApoE were linked to SDMD, FE-SDMD and subclinical MetS. An additional clinical concern for patients with SDMD is the potential interaction between SDMD and subclinical MetS, which can result in decreased HDLc and RCT levels and increased ApoB/ApoA ratio, when subclinical MetS develops. Thus, SDMD appears to be a pre-proatherogenic state, because of decreased LCAT and increased free cholesterol (see below) and its intersections with subclinical MetS.

### LCAT as a key factor in SDMD and MDMD

An apparent distinction between the present investigation and that of Maes et al. [35] is that the latter did not document reduced LCAT levels in individuals with depression. In contrast, the current study identified LCAT as the most significant predictor of the phenome of depression, with free cholesterol ranking second. To clarify these potential distinctions, we reexamined the data set from Maes et al. [35] to decipher the effects of LCAT on the phenome more precisely through mediation analysis. In doing so, both investigations demonstrated that LCAT exerts substantial impacts on the phenome that are entirely mediated by either atherogenicity [35] or free cholesterol and brooding (as in this study). Thus, LCAT influences the phenotype as measured is FE-SDMD and more severe depression, which is distinguished by an elevated ROI index.

This finding is highly intriguing as it demonstrates that decreased LCAT, which results in elevated free cholesterol (and consequently its detrimental impacts; see below), is the critical determinant in FE-SDMD, albeit without the complete manifestation of atherogenicity. Indeed, FE-SDMD exhibits characteristics of a pre-proatherogenic state because of reduced LCAT and elevated free cholesterol. Nonetheless, as the disease progresses (with an increasing ROI) and more relapses occur, the effect of atherogenicity increases, due to reduced RCT and elevated free cholesterol and atherogenicity, as well as intersections between SDMD and subclinical MetS that may develop into MetS. These findings may suggest that SDMD is a pre-proatherogenic state with a pathophysiology that is different from what is observed in subclinical MetS. As such, SDMD may well constitute an independent risk factor for atherogenicity thereby contributing to MetS.

### LCAT, brooding and the phenome of depression

As previously mentioned [34], a portion of the variability in the severity of the SDMD phenome can be attributed to the cognitive process of brooding (rumination over a variety of negative factors) [63]. Like Maes et al. [34], we have accounted for brooding in our analyses to estimate the relationships between lipid fractions and the portion of the phenome that is not influenced by brooding. By doing so, we discerned that these residualized phenome scores were significantly correlated with free cholesterol, which in fact mediated the effects of LCAT on the phenome. Therefore, SDMD manifests as a dual-component syndrome, consisting of lipid-associated pathways, and a brooming or pondering component which is partly predicted by the same lipid pathways. In alternative formulations, the severity of illness and suicidal behaviors in SDMD and FE-SDMD are determined through the synergistic influence of lipid-associated characteristics and cognitive features, which are partially influenced by the same lipids. In a prior study, we demonstrated a comparable correlation between immune variables and observed outcomes: we found that immune pathways accounted for a fraction of the variability in brooding, whereas both the immune pathways and brooding accounted for a substantial portion of the variance in the phenome [34]. Thus, the combined influence of lipid-associated and immune pathways may account for a greater proportion of the variability observed in rumination and the phenome of depression. Further research should investigate whether the clinical depression data reveal combined effects of both pathway types.

### Mechanistic importance of LCAT in the pathophysiology of depression

LCAT has the most significant impact on the severity of the FE-SDMD phenotype (this study) and a significant mediated effects on recurrent depression as well, as mentioned previously [35]. Reduced LCAT is a crucial factor in HDLc metabolism and, as previously mentioned, may be the driving force behind RCT [1, 27]. Nevertheless, the current study failed to identify any significant reductions in RCT in SDMD or FE-SDMD, although those patients exhibited decreased LCAT. Conversely, very low RCT levels are identified in recurrent depression [35]. Based on these correlations and the observed impacts of reduced LCAT on the phenotype [35], this study), we may propose that decreased LCAT could serve as a catalyst for the progression of atherogenicity and the escalating ROI.

The significance of LCAT for atherogenicity remains a subject of intense debate, despite the passage of four to five decades of research [22, 51]. Numerous large-scale studies failed to identify the association between a decreased LCAT and an increased cardiovascular risk. Conversely, there is a clear association between LCAT deficiency and compromised HDL maturation, as well as accumulation of putative preβ-HDL and upregulation of ApoE-rich HDL particle expression [22]. Research on rodents and primates indicates that LCAT has atheroprotective properties [51]. Additionally, LCAT has the potential to impact the vulnerability to atherosclerosis by modulating LDL receptors and, by extension, LDL metabolism [9]. It is noteworthy to mention that recombinant human LCAT treatment might exhibit some degree of effectiveness in the management of atherosclerosis [66].

While the precise mechanism by which LCAT contributes to atherogenicity remains unknown, LCAT plays a significant role as an antioxidant enzyme [64]. LCAT remains active for a duration of 10 hours when exposed to mild free-radical generators. Therefore, LCAT can impede the oxidation process of LDL, potentially hinder the buildup of oxidized lipids in LDL, and contribute to the hydrolysis of oxidized lipoproteins [18, 64]. Furthermore, Hine et al. [18] found that the combination of LCAT, ApoA1, and PON1 extends the time during which HDL can inhibit LDL oxidation. Decreased LCAT levels may therefore contribute to the elevated oxidative stress that has been identified in depressive disorders [28]. Patients diagnosed with SDMD exhibit elevated levels of malondialdehyde (MDA) in their serum, according to recent findings (Maes et al., to be submitted). Theoretically, the resulting surge in oxidative stress could potentially contribute to elevated levels of cholesterol oxidation and, consequently, oxysterols, which have been implicated in the development of certain neurodegenerative disorders, including Alzheimer’s disease [15].

Furthermore, decreased LCAT diminishes the ability of immature HDL to neutralize LPS in rodents, thus increasing the inflammation induced by LPS [44]. Significantly, MDD is associated with elevated LPS load in peripheral blood, which may promote inflammation and oxidative stress, including increased LDL oxidation and IgG responses to oxidized LDL, both of which are crucial pathways in the development of atherosclerosis [29]. Consequently, the decreased activity of the LCAT enzyme in SDMD may be linked to an increase in LDL oxidation, which may result in the development of atherosclerosis. Furthermore, oxysterols have the potential to induce pro-inflammatory, cytotoxic, and pro-apoptotic responses [42]. As such, lowered LCAT in peripheral blood may contribute to the immune-inflammatory and oxidative stress pathophysiology of depression [27–29].

### Effects of increased free cholesterol and ApoE in SDMD

As mentioned in the introduction, an elevation in free cholesterol levels can potentially cause cytotoxicity to macrophages and cell membranes, thus playing a role in the development of atherosclerosis [58, 59]. Extensive literature exists regarding the neurotoxic and cytotoxic consequences of hypercholesterolemia. Significantly, cholesterol-induced macrophage mortality can be induced by free cholesterol via the induction of the unfolded protein response and the depletion of endoplasmic reticulum calcium stores [67]. Cholesterol has the potential to induce lipid peroxidation and cellular demise in liver cells [49]. Elevated levels of free cholesterol therefore promote degenerative harm to peripheral tissues, including endothelial cells.

Moreover, the elevation in ApoE levels that we detected in our research could potentially contribute to peripheral abnormalities. Robust correlations were observed between ApoE and triglycerides, ApoB and free cholesterol, and total cholesterol in the sample used in this study. The correlation with triglycerides was particularly strong (r=0.514, p<0.001). Plasma apoE is understood to regulate the metabolism of triglyceride-rich lipoproteins and may account for 20– 40% of the variability in triglyceride levels in humans [31, 32]. This finding is consistent with our own results, which indicate a shared variance of approximately 25%. Triglyceridemia can be induced by the accumulation of ApoE, which inhibits the lipolysis of very low-density lipoprotein triglyceride particles and stimulates their production [31, 32]. Conversely, a diminished ApoE level could impede the clearance of triglyceride-rich lipoproteins from the plasma. As a result, the elevated ApoE levels observed in SDMD may potentially contribute to the pre-proatherogenic state characterized by decreased LCAT and elevated free cholesterol.

### Are there any central effects of peripheral changes in lipid metabolism

Another unresolved inquiry pertains to the potential ramifications of the fluctuations in serum LCAT activity on the brain. LCAT is produced endogenously in the brain, particularly by astrocytes, suggesting that it might have a significant impact on the development of brain lipoproteins and nascent lipoproteins derived from glial cells [19]. Additionally, brain LCAT esterifies cholesterol on apoE-lipoproteins derived from glial cells. Should a genetic element be present in the relationship between depression and decreased LCAT [27], it is possible that decreased brain LCAT would also be associated with depression, leading to significant implications for cholesterol homeostasis and metabolism in the brain. Consequently, this could result in neuronal damage induced by the direct impacts of toxic lipids or their oxidatively derived metabolites. For example, the aggregation of amyloid β (Aβ) on lipid bilayers is expedited through the production of larger aggregates in the presence of free cholesterol [17]. Furthermore, the surface dissociation of these aggregates is accelerated by free cholesterol. The development of AD may be influenced by hypercholesterolemia in old age and hypercholesterolemia in middle age [11].

BBB permeability, astrogliosis, and neurocognitive deficits may result from elevated circulating cholesterol levels caused by a high-cholesterol diet [14]. In addition, unlike cholesterol, certain byproducts of cholesterol oxidation may cross the blood-brain barrier (BBB) to stimulate microglia and cause neurotoxic effects in the brain, establishing a link between hypercholesterolemia and conditions such as Alzheimer’s disease [7, 48, 54]. Furthermore, albumin-bound non-esterified fatty acids may traverse the BBB via passive diffusion [12, 45]. Overall, elevated levels of free cholesterol may promote central degenerative disorders, such as by impairing the blood-brain barrier (BBB) and facilitating the formation of oxysterols.

Furthermore, it is worth noting that ApoE is produced through cellular synthesis in neurons and is susceptible to various stimuli, including oxidative stress, trauma, and advancing age [31]. Neuronal ApoE synthesis may result in neuropathology, particularly in the formation of ApoE4 products [31].

## Limitations

The potential significance of the present investigation might have been further enhanced had oxysterols, omega-3 polyunsaturated fatty acids, and vitamin E been quantified. The correlation between LCAT polymorphisms and SDMD and MDMD warrants further investigation, as certain LCAT gene variants are linked to HDL metabolism, exacerbation of dyslipidemia, and acute coronary syndrome [5, 62]. In the transition from FE-SDMD to new episodes of SDMD and MDMD, lipid metabolism should be the subject of prospective studies. This research should aim to identify the specific lipid disorders that contribute to increased atherogenicity and the development of MetS in depression.

## Conclusions

Free cholesterol and ApoE levels are elevated in subjects with SDMD and FE-SDMD relative to healthy controls, while LCAT levels are decreased. In both SDMD and FE-SDMD, no substantial changes were observed in HDLc, ApoA1, RCT, and ApoB. These results further show that SDMD and MDMD are distinct subgroups of MDD which are differentiated by their lipid (and neuro-immune) profiles. Significant effects of LCAT on the manifestation of depression, including suicidal behaviors, are completely mediated by free cholesterol and brooding.

Compared to healthy controls, SDMD and FE-SDMD patients without any indication of subclinical MetS have decreased LCAT and elevated free cholesterol levels. These two biomarkers are not significantly associated with subclinical MetS and are, therefore, unique biomarkers of FE-SDMD and SDMD. Moreover, significant interactions exist between the SDMD diagnosis and subclinical MetS in SDMD and FE-SDMD, leading to a reduction in HDLc and RCT levels, as well as an increase in the ApoB/ApoA ratio.

FE-SDMD and SDMD are pre-proatherogenic states due to their intersections with subclinical MetS, decreased LCAT, and increased free cholesterol and ApoE. The activation of neuro-oxidative and neuro-immune pathways may be induced by these abnormalities. Therefore, we believe that it is necessary for all persons with FE-SDMD to undertake lipid screening using the assays for free cholesterol, LCAT, and ApoE. As such, it is possible to identify individuals who are at risk of developing neuroimmune and neuro-oxidative disorders, increased atherogenicity, and MetS and more severe forms of recurrent depression. Consequently, a preventive therapy may be initiated targeting lowered LCAT and increased free cholesterol levels.

## Ethical approval and consent to participate

The research project (IRB no.351/63) was approved by the Institutional Review Board of Chulalongkorn University’s institutional ethics board, Bangkok, Thailand, which follows the International Guideline for Human Research protection as required by the Declaration of Helsinki, The Belmont Report, CIOMS Guideline and International Conference on Harmonization in Good Clinical Practice (ICH-GCP). All participants signed the appropriate institutional informed consent forms before data collection.

## Availability of data and materials

MM will reply to reasonable requests for the dataset used in the current study after it has been fully utilized by all authors.

## Credit author’s contributions

AV and MM conducted the current study’s design. The data was gathered by AV. The statistical evaluation was performed by MM. MM wrote the first draft. All authors contributed to the editing of the work, and they have all given their consent for submission of the completed version.

## Acknowledgments

Not applicable.

