## supplementary file for "First-episode mild depression in young adults is a pre-proatherogenic condition even in the absence of subclinical metabolic syndrome: lowered lecithin-cholesterol acyltransferase as a key factor"

**ELECTRONIC SUPPLEMENTARY FILE**

**ESF, Table 1** Raw metabolic data in students with simple dysmood disorder (1) and normal control students (0).

| **Estimates** | | | | | |
| --- | --- | --- | --- | --- | --- |
| Dependent Variable | SDMD_HC | Mean | Std. Error | 95% Confidence Interval | |
|  |  |  |  | Lower Bound | Upper Bound |
| Total cholesterol (c) | HC | 195.521^a^ | 5.056 | 185.494 | 205.549 |
|  | SDMD | 206.139^a^ | 4.181 | 197.846 | 214.431 |
| HDLc | HC | 60.648^a^ | 1.728 | 57.221 | 64.075 |
|  | SDMD | 61.232^a^ | 1.429 | 58.399 | 64.066 |
| Triglycerides | HC | 80.224^a^ | 9.574 | 61.236 | 99.212 |
|  | SDMD | 95.759^a^ | 7.917 | 80.057 | 111.461 |
| LDLc | HC | 118.831^a^ | 4.194 | 110.514 | 127.148 |
|  | SDMD | 125.800^a^ | 3.468 | 118.922 | 132.678 |
| Free cholesterol | HC | 37.444^a^ | 1.252 | 34.961 | 39.927 |
|  | SDMD | 45.307^a^ | 1.035 | 43.253 | 47.360 |
| ApoA1 | HC | 138.845^a^ | 3.722 | 131.463 | 146.227 |
|  | SDMD | 142.056^a^ | 3.078 | 135.951 | 148.161 |
| ApoB | HC | 81.092^a^ | 2.661 | 75.815 | 86.370 |
|  | SDMD | 87.261^a^ | 2.200 | 82.897 | 91.625 |
| ApoE (ng/mL) | HC | 7695.369^a^ | 784.015 | 6140.459 | 9250.278 |
|  | SDMD | 11069.633^a^ | 648.347 | 9783.789 | 12355.477 |
| a. Covariates appearing in the model are evaluated at the following values: Age = 22.8678, Sex = .16, BMI = 22.3801. All data in mg/dL, except ApoE. | | | | | |

| **Univariate Tests** | | | | | | |
| --- | --- | --- | --- | --- | --- | --- |
| Dependent Variable | | Sum of Squares | df | Mean Square | F | Sig. |
| Total cholesterol | Contrast | 2856.249 | 1 | 2856.249 | 2.583 | .111 |
|  | Error | 113892.212 | 103 | 1105.750 |  |  |
| HDLc | Contrast | 8.648 | 1 | 8.648 | .067 | .796 |
|  | Error | 13300.343 | 103 | 129.130 |  |  |
| Triglycerides | Contrast | 6114.400 | 1 | 6114.400 | 1.542 | .217 |
|  | Error | 408376.666 | 103 | 3964.822 |  |  |
| LDLs | Contrast | 1230.617 | 1 | 1230.617 | 1.618 | .206 |
|  | Error | 78353.357 | 103 | 760.712 |  |  |
| Free cholesterol | Contrast | 1566.292 | 1 | 1566.292 | 23.104 | .000 |
|  | Error | 6982.772 | 103 | 67.794 |  |  |
| ApoA1 | Contrast | 261.282 | 1 | 261.282 | .436 | .511 |
|  | Error | 61724.886 | 103 | 599.271 |  |  |
| ApoB | Contrast | 964.060 | 1 | 964.060 | 3.148 | .079 |
|  | Error | 31544.758 | 103 | 306.260 |  |  |
| ApoE | Contrast | 288478485.989 | 1 | 288478485.989 | 10.850 | .001 |
|  | Error | 2738520251.951 | 103 | 26587575.262 |  |  |
| The F tests the effect of SDMD_HC. This test is based on the linearly independent pairwise comparisons among the estimated marginal means. | | | | | | |

**ESF, Table 2**. Lipid measurements in participants divided into different stages of subclinical metabolic syndrome, namely stage 0, stage 1 and stage 2.

| **Estimates** | | | | | |
| --- | --- | --- | --- | --- | --- |
| Dependent Variable | METs score 3 groups | Mean | Std. Error | 95% Confidence Interval | |
|  |  |  |  | Lower Bound | Upper Bound |
| Total cholesterol (c) | 0 | .082^a^ | .134 | -.185 | .348 |
|  | 1 | -.245^a^ | .222 | -.685 | .196 |
|  | 2 | -.112^a^ | .356 | -.819 | .594 |
| HDLc | 0 | .295^a^ | .117 | .062 | .528 |
|  | 1 | -.335^a^ | .194 | -.720 | .050 |
|  | 2 | -.956^a^ | .311 | -1.573 | -.338 |
| Triglycerides | 0 | -.228^a^ | .132 | -.491 | .034 |
|  | 1 | .061^a^ | .219 | -.374 | .495 |
|  | 2 | 1.135^a^ | .351 | .438 | 1.831 |
| LDLc | 0 | .136^a^ | .137 | -.137 | .408 |
|  | 1 | -.229^a^ | .227 | -.679 | .221 |
|  | 2 | -.267^a^ | .364 | -.989 | .455 |
| Free cholesterol | 0 | .037^a^ | .122 | -.205 | .279 |
|  | 1 | -.228^a^ | .201 | -.628 | .171 |
|  | 2 | -.306^a^ | .323 | -.947 | .335 |
| ApoA1 | 0 | .159^a^ | .132 | -.104 | .421 |
|  | 1 | -.248^a^ | .218 | -.681 | .185 |
|  | 2 | -.706^a^ | .350 | -1.401 | -.011 |
| ApoB | 0 | -.104^a^ | .128 | -.357 | .150 |
|  | 1 | -.085^a^ | .211 | -.505 | .334 |
|  | 2 | .513^a^ | .339 | -.160 | 1.185 |
| ApoE | 0 | -.297^a^ | .109 | -.513 | -.081 |
|  | 1 | .171^a^ | .180 | -.186 | .528 |
|  | 2 | .955^a^ | .289 | .382 | 1.528 |
| LCAT | 0 | .135^a^ | .126 | -.115 | .386 |
|  | 1 | -.001^a^ | .208 | -.415 | .413 |
|  | 2 | .173^a^ | .334 | -.491 | .837 |
| ApoB/ApoA | 0 | -.183^a^ | .113 | -.408 | .041 |
|  | 1 | .092^a^ | .187 | -.278 | .463 |
|  | 2 | .736^a^ | .300 | .141 | 1.330 |
| RCT | 0 | .269^a^ | .123 | .025 | .512 |
|  | 1 | -.259^a^ | .203 | -.661 | .143 |
|  | 2 | -.663^a^ | .325 | -1.309 | -.018 |
| Atherogenicity | 0 | -.109^a^ | .121 | -.349 | .131 |
|  | 1 | -.067^a^ | .200 | -.463 | .329 |
|  | 2 | .605^a^ | .320 | -.031 | 1.241 |
| Athero/LCAT | 0 | -.090^a^ | .114 | -.316 | .136 |
|  | 1 | -.032^a^ | .188 | -.405 | .342 |
|  | 2 | .251^a^ | .302 | -.348 | .850 |
| Athero/RCT | 0 | -.262^a^ | .109 | -.479 | -.045 |
|  | 1 | .150^a^ | .181 | -.208 | .509 |
|  | 2 | .929^a^ | .290 | .354 | 1.505 |
| a. Covariates appearing in the model are evaluated at the following values: Age = 22.9108, Sex = .15, BMI = 22.4290. All data are shown in z scores. | | | | | |

| **Univariate Tests** | | | | | | |
| --- | --- | --- | --- | --- | --- | --- |
| Dependent Variable | | Sum of Squares | df | Mean Square | F | Sig. |
| Total cholesterol (c) | Contrast | 1.261 | 2 | .631 | .686 | .506 |
|  | Error | 87.289 | 95 | .919 |  |  |
| HDLc | Contrast | 9.367 | 2 | 4.684 | 6.673 | .002 |
|  | Error | 66.685 | 95 | .702 |  |  |
| Triglycerides | Contrast | 10.218 | 2 | 5.109 | 5.720 | .005 |
|  | Error | 84.848 | 95 | .893 |  |  |
| LDLc | Contrast | 1.750 | 2 | .875 | .912 | .405 |
|  | Error | 91.176 | 95 | .960 |  |  |
| Free cholesterol | Contrast | 1.020 | 2 | .510 | .675 | .512 |
|  | Error | 71.829 | 95 | .756 |  |  |
| ApoA1 | Contrast | 4.337 | 2 | 2.168 | 2.440 | .093 |
|  | Error | 84.409 | 95 | .889 |  |  |
| ApoB | Contrast | 2.459 | 2 | 1.229 | 1.477 | .234 |
|  | Error | 79.066 | 95 | .832 |  |  |
| ApoE | Contrast | 8.560 | 2 | 4.280 | 7.086 | .001 |
|  | Error | 57.378 | 95 | .604 |  |  |
| LCAT | Contrast | .330 | 2 | .165 | .203 | .816 |
|  | Error | 77.075 | 95 | .811 |  |  |
| ApoB/ApoA | Contrast | 4.544 | 2 | 2.272 | 3.489 | .034 |
|  | Error | 61.861 | 95 | .651 |  |  |
| RCT | Contrast | 5.580 | 2 | 2.790 | 3.638 | .030 |
|  | Error | 72.861 | 95 | .767 |  |  |
| Atherogenicity | Contrast | 3.181 | 2 | 1.590 | 2.137 | .124 |
|  | Error | 70.692 | 95 | .744 |  |  |
| Athero/LCAT | Contrast | .653 | 2 | .326 | .494 | .612 |
|  | Error | 62.746 | 95 | .660 |  |  |
| Athero/RCT | Contrast | 7.680 | 2 | 3.840 | 6.300 | .003 |
|  | Error | 57.906 | 95 | .610 |  |  |
| The F tests the effect of METs score 3 groups. This test is based on the linearly independent pairwise comparisons among the estimated marginal means. l | | | | | | |

| **Pairwise Comparisons** | | | | | | | |
| --- | --- | --- | --- | --- | --- | --- | --- |
| Dependent Variable | (I) METs score 3 groups | (J) METs score 3 groups | Mean Difference (I-J) | Std. Error | Sig.^b^ | 95% Confidence Interval for Difference^b^ | |
|  |  |  |  |  |  | Lower Bound | Upper Bound |
| Total cholesterol (c) | 0 | 1 | .326 | .279 | .245 | -.227 | .880 |
|  |  | 2 | .194 | .413 | .640 | -.626 | 1.014 |
|  | 1 | 0 | -.326 | .279 | .245 | -.880 | .227 |
|  |  | 2 | -.132 | .382 | .730 | -.890 | .626 |
|  | 2 | 0 | -.194 | .413 | .640 | -1.014 | .626 |
|  |  | 1 | .132 | .382 | .730 | -.626 | .890 |
| HDLc | 0 | 1 | .630^*^ | .244 | .011 | .146 | 1.114 |
|  |  | 2 | 1.250^*^ | .361 | .001 | .534 | 1.967 |
|  | 1 | 0 | -.630^*^ | .244 | .011 | -1.114 | -.146 |
|  |  | 2 | .620 | .334 | .066 | -.042 | 1.283 |
|  | 2 | 0 | -1.250^*^ | .361 | .001 | -1.967 | -.534 |
|  |  | 1 | -.620 | .334 | .066 | -1.283 | .042 |
| Triglycerides | 0 | 1 | -.289 | .275 | .296 | -.835 | .257 |
|  |  | 2 | -1.363^*^ | .407 | .001 | -2.172 | -.555 |
|  | 1 | 0 | .289 | .275 | .296 | -.257 | .835 |
|  |  | 2 | -1.074^*^ | .376 | .005 | -1.821 | -.327 |
|  | 2 | 0 | 1.363^*^ | .407 | .001 | .555 | 2.172 |
|  |  | 1 | 1.074^*^ | .376 | .005 | .327 | 1.821 |
| LDLc | 0 | 1 | .365 | .285 | .203 | -.201 | .931 |
|  |  | 2 | .403 | .422 | .343 | -.436 | 1.241 |
|  | 1 | 0 | -.365 | .285 | .203 | -.931 | .201 |
|  |  | 2 | .038 | .390 | .924 | -.737 | .812 |
|  | 2 | 0 | -.403 | .422 | .343 | -1.241 | .436 |
|  |  | 1 | -.038 | .390 | .924 | -.812 | .737 |
| Free cholesterol | 0 | 1 | .265 | .253 | .297 | -.237 | .767 |
|  |  | 2 | .343 | .375 | .362 | -.401 | 1.087 |
|  | 1 | 0 | -.265 | .253 | .297 | -.767 | .237 |
|  |  | 2 | .078 | .346 | .822 | -.610 | .765 |
|  | 2 | 0 | -.343 | .375 | .362 | -1.087 | .401 |
|  |  | 1 | -.078 | .346 | .822 | -.765 | .610 |
| ApoA1 | 0 | 1 | .406 | .274 | .142 | -.138 | .951 |
|  |  | 2 | .864^*^ | .406 | .036 | .058 | 1.671 |
|  | 1 | 0 | -.406 | .274 | .142 | -.951 | .138 |
|  |  | 2 | .458 | .375 | .226 | -.287 | 1.203 |
|  | 2 | 0 | -.864^*^ | .406 | .036 | -1.671 | -.058 |
|  |  | 1 | -.458 | .375 | .226 | -1.203 | .287 |
| ApoB | 0 | 1 | -.018 | .265 | .945 | -.545 | .508 |
|  |  | 2 | -.617 | .393 | .120 | -1.397 | .164 |
|  | 1 | 0 | .018 | .265 | .945 | -.508 | .545 |
|  |  | 2 | -.598 | .363 | .103 | -1.320 | .123 |
|  | 2 | 0 | .617 | .393 | .120 | -.164 | 1.397 |
|  |  | 1 | .598 | .363 | .103 | -.123 | 1.320 |
| ApoE | 0 | 1 | -.468^*^ | .226 | .041 | -.917 | -.019 |
|  |  | 2 | -1.252^*^ | .335 | .000 | -1.917 | -.587 |
|  | 1 | 0 | .468^*^ | .226 | .041 | .019 | .917 |
|  |  | 2 | -.784^*^ | .310 | .013 | -1.399 | -.170 |
|  | 2 | 0 | 1.252^*^ | .335 | .000 | .587 | 1.917 |
|  |  | 1 | .784^*^ | .310 | .013 | .170 | 1.399 |
| LCAT | 0 | 1 | .137 | .262 | .603 | -.384 | .657 |
|  |  | 2 | -.038 | .388 | .923 | -.808 | .733 |
|  | 1 | 0 | -.137 | .262 | .603 | -.657 | .384 |
|  |  | 2 | -.174 | .359 | .629 | -.886 | .538 |
|  | 2 | 0 | .038 | .388 | .923 | -.733 | .808 |
|  |  | 1 | .174 | .359 | .629 | -.538 | .886 |
| ApoB/ApoA | 0 | 1 | -.276 | .235 | .243 | -.742 | .190 |
|  |  | 2 | -.919^*^ | .348 | .010 | -1.609 | -.228 |
|  | 1 | 0 | .276 | .235 | .243 | -.190 | .742 |
|  |  | 2 | -.643^*^ | .321 | .048 | -1.281 | -.005 |
|  | 2 | 0 | .919^*^ | .348 | .010 | .228 | 1.609 |
|  |  | 1 | .643^*^ | .321 | .048 | .005 | 1.281 |
| RCT | 0 | 1 | .528^*^ | .255 | .041 | .022 | 1.033 |
|  |  | 2 | .932^*^ | .378 | .015 | .183 | 1.682 |
|  | 1 | 0 | -.528^*^ | .255 | .041 | -1.033 | -.022 |
|  |  | 2 | .404 | .349 | .249 | -.288 | 1.097 |
|  | 2 | 0 | -.932^*^ | .378 | .015 | -1.682 | -.183 |
|  |  | 1 | -.404 | .349 | .249 | -1.097 | .288 |
| Atherogenicity | 0 | 1 | -.042 | .251 | .868 | -.540 | .456 |
|  |  | 2 | -.714 | .372 | .058 | -1.452 | .025 |
|  | 1 | 0 | .042 | .251 | .868 | -.456 | .540 |
|  |  | 2 | -.672 | .344 | .053 | -1.354 | .010 |
|  | 2 | 0 | .714 | .372 | .058 | -.025 | 1.452 |
|  |  | 1 | .672 | .344 | .053 | -.010 | 1.354 |
| Athero/LCAT | 0 | 1 | -.058 | .236 | .807 | -.527 | .411 |
|  |  | 2 | -.340 | .350 | .334 | -1.036 | .355 |
|  | 1 | 0 | .058 | .236 | .807 | -.411 | .527 |
|  |  | 2 | -.282 | .324 | .385 | -.925 | .360 |
|  | 2 | 0 | .340 | .350 | .334 | -.355 | 1.036 |
|  |  | 1 | .282 | .324 | .385 | -.360 | .925 |
| Athero/RCT | 0 | 1 | -.412 | .227 | .073 | -.863 | .039 |
|  |  | 2 | -1.191^*^ | .337 | .001 | -1.859 | -.523 |
|  | 1 | 0 | .412 | .227 | .073 | -.039 | .863 |
|  |  | 2 | -.779^*^ | .311 | .014 | -1.396 | -.162 |
|  | 2 | 0 | 1.191^*^ | .337 | .001 | .523 | 1.859 |
|  |  | 1 | .779^*^ | .311 | .014 | .162 | 1.396 |
| Based on estimated marginal means | | | | | | | |
| *. The mean difference is significant at the .05 level. | | | | | | | |
| b. Adjustment for multiple comparisons: Least Significant Difference (equivalent to no adjustments). | | | | | | | |

**ESF Table 3** Interactions between the clinical diagnosis simple dysmood disorder (SDMD) versus control (SDMD_HC) X the three stages (0, 1 and 2) of subclinical metabolic syndrome (METs_Score012).

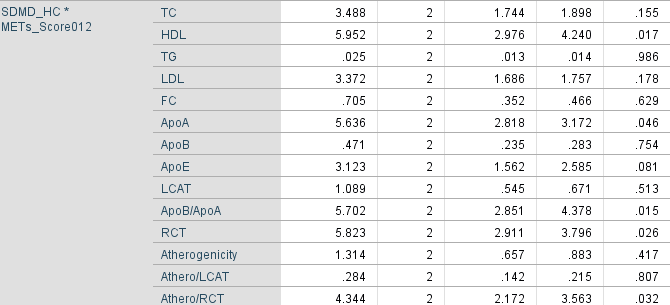

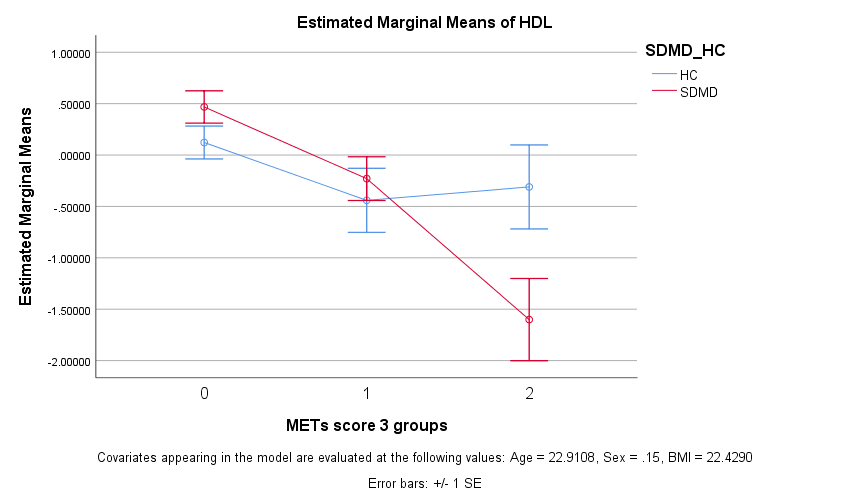

**ESF, Figure 1** Serum high density lipoprotein cholesterol (HDL) levels in students with simple dysmood disorder (SDMD) and healthy control students (HC) and in the three subclinical metabolic syndrome (METs) stages (stages 0, 1 and 2). The interaction pattern was significant (see ESF, Table 3). Values are shown as z scores (SE).

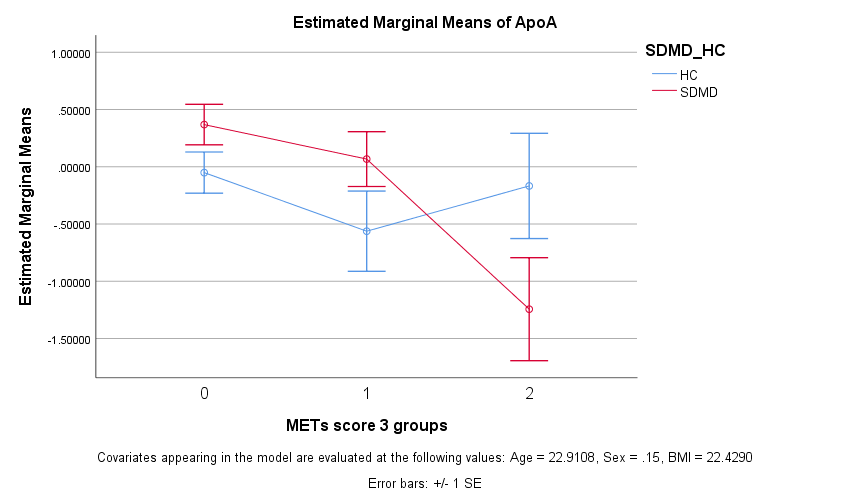

**ESF, Figure 2** Serum apolipoprotein A1 (ApoA) levels in students with simple dysmood disorder (SDMD) and healthy control students (HC) and in the three subclinical metabolic syndrome stages (stages 0, 1 and 2). The interaction pattern was significant (see ESF, Table 3). Values are shown as z scores (SE).

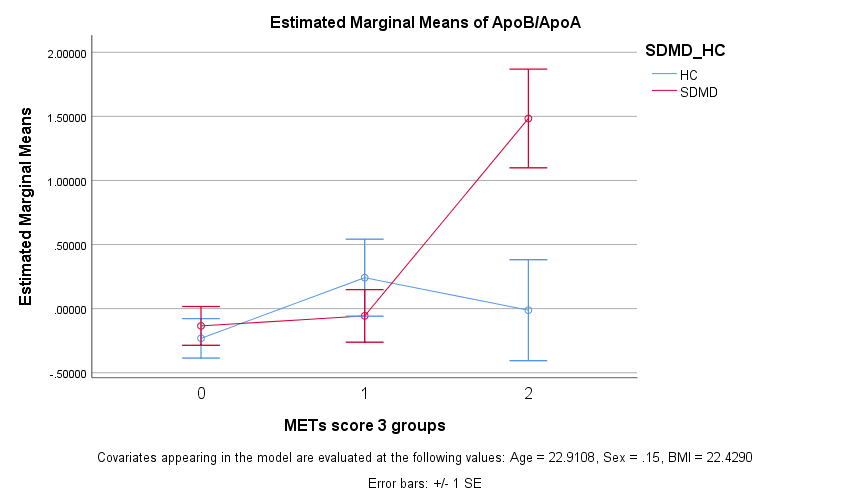

**ESF, Figure 3** The ApoB/ApoA1 ratio in students with simple dysmood disorder (SDMD) and healthy control students (HC) and in the three subclinical metabolic syndrome stages (stages 0, 1 and 2). The interaction pattern was significant (see ESF, Table 3). Values are shown as z scores (SE).

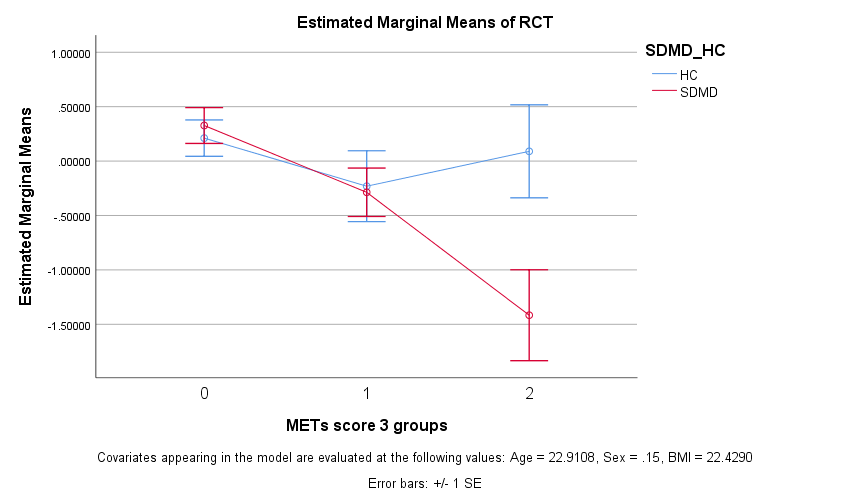

**ESF, Figure 4** Reverse cholesterol transport (RCT) index in students with simple dysmood disorder (SDMD) and healthy control students (HC) and in the three subclinical metabolic syndrome stages (stages 0, 1 and 2). The interaction pattern was significant (see ESF, Table 3). Values are shown as z scores (SE).

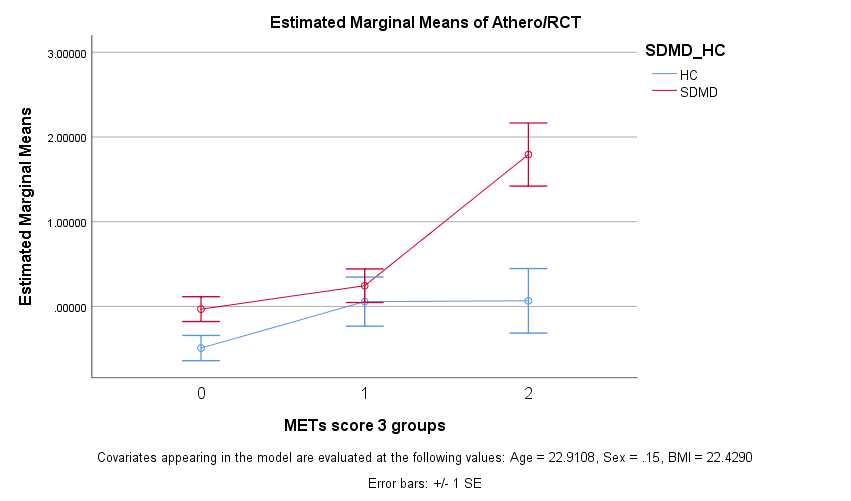

**ESF, Figure 5** The atherogenicity / reverse cholesterol transport (RCT) ratio in students with simple dysmood disorder (SDMD) and healthy control students (HC) and in the three subclinical metabolic syndrome stages (stages 0, 1 and 2). The interaction pattern was significant (see ESF, Table 3). Values are shown as z scores (SE).
